## Supplemental Table 1 for "Brain connectivity changes underlying depression and fatigue in relapsing-remitting multiple sclerosis: a systematic review"

S1 Table. An overview of all studies read in full and final decision.

| Author | Year | Title | Decision |
|---|---|---|---|
| A. Alshehri; O. Al-iedani; J. Arm; N. Gholizadeh; T. Billiet; R. Lea; J. Lechner-Scott; S. Ramadan | 2022 | Neural diffusion tensor imaging metrics correlate with clinical measures in people with relapsing-remitting MS | Final Inclusion |
| A. Altermatt; L. Gaetano; S. Magon; D. A. Häring; D. Tomic; J. Wuerfel; E. W. Radue; L. Kappos; T. Sprenger | 2018 | Clinical Correlations of Brain Lesion Location in Multiple Sclerosis: Voxel-Based Analysis of a Large Clinical Trial Dataset | Final Inclusion |
| A. Andravizou; V. Siokas; A. Artemiadis; C. Bakirtzis; A. M. Aloizou; N. Grigoriadis; M. H. Kosmidis; G. Nasios; L. Messinis; G. Hadjigeorgiou; E. Dardiotis; E. Peristeri | 2020 | Clinically reliable cognitive decline in relapsing remitting multiple sclerosis: Is it the tip of the iceberg? | no D/F |
| A. Bisecco; G. Caiazzo; A. d'Ambrosio; R. Sacco; S. Bonavita; R. Docimo; M. Cirillo; E. Pagani; M. Filippi; F. Esposito; G. Tedeschi; A. Gallo | 2016 | Fatigue in multiple sclerosis: The contribution of occult white matter damage | Final Inclusion |
| A. C. Vogel; H. Schmidt; S. Loud; R. McBurney; F. J. Mateen | 2020 | Impact of the COVID-19 pandemic on the health care of >1,000 People living with multiple sclerosis: A cross-sectional study | do not correlate MRI to D/F |
| A. Carotenuto; H. Wilson; B. Giordano; S. P. Caminiti; Z. Chappell; S. C. R. Williams; A. Hammers; E. Silber; P. Brex; M. Politis | 2020 | Impaired connectivity within neuromodulatory networks in multiple sclerosis and clinical implications | Final Inclusion |
| A. Conte; C. Giannì; D. Belvisi; A. Cortese; N. Petsas; M. Tartaglia; P. Cimino; E. Millefiorini; A. Berardelli; P. Pantano | 2020 | Deep grey matter involvement and altered sensory gating in multiple sclerosis | no D/F |
| A. Damasceno; B. P. Damasceno; F. Cendes | 2016 | Atrophy of reward-related striatal structures in fatigued MS patients is independent of physical disability | Final Inclusion |
| A. Damasceno; L. R. Pimentel-Silva; B. P. Damasceno; F. Cendes | 2020 | Cognitive trajectories in relapsing–remitting multiple sclerosis: A longitudinal 6-year study | no D/F |
| A. E. Williams; J. T. Vietri; G. Isherwood; A. Flor | 2014 | Symptoms and Association with Health Outcomes in Relapsing-Remitting Multiple Sclerosis: Results of a US Patient Survey | no MRI |
| A. Ernst; M. Sourty; D. Roquet; V. Noblet; D. Gounot; F. Blanc; J. De Seze; L. Manning | 2016 | Functional and structural cerebral changes in key brain regions after a facilitation programme for episodic future thought in relapsing-remitting multiple sclerosis patients | do not correlate MRI to D/F |
| A. Ernst; M. Sourty; D. Roquet; V. Noblet; D. Gounot; F. Blanc; J. de Seze; L. Manning | 2018 | Benefits from an autobiographical memory facilitation programme in relapsing-remitting multiple sclerosis patients: a clinical and neuroimaging study | do not correlate MRI to D/F |
| A. Ernst; V. Noblet; E. Denkova; F. Blanc; J. de Seze; D. Gounot; L. Manning | 2015 | Functional cerebral changes in multiple sclerosis patients during an autobiographical memory test | Sample size <20 |
| A. Harel; D. Sperling; M. Petracca; A. Ntranos; I. Katz-Sand; S. Krieger; F. Lublin; Z. Wang; Y. Liu; M. Inglese | 2018 | Brain microstructural injury occurs in patients with RRMS despite €no evidence of disease activity' | wrong D/F test |

| | | | |
|---|---|---|---|
| A. J. Cruz Gomez; N. V. Campos; A. Belenguer; C. Avila; C. Forn | 2013 | Regional Brain Atrophy and Functional Connectivity Changes Related to Fatigue in Multiple Sclerosis | Final Inclusion |
| A. K. Andreasen; J. Jakobsen; L. Soerensen; H. Andersen; T. Petersen; C. R. Bjarkam; J. Ahdidan | 2010 | Regional brain atrophy in primary fatigued patients with multiple sclerosis | Final Inclusion |
| A. Kever; K. Buyukturkoglu; S. N. Levin; C. S. Riley; P. De Jager; V. M. Leavitt | 2022 | Associations of social network structure with cognition and amygdala volume in multiple sclerosis: An exploratory investigation | Final Inclusion |
| A. L. Ruiz-Rizzo; P. Bublak; S. Kluckow; K. Finke; C. Gaser; M. Schwab; D. Güllmar; H. J. Müller; O. Witte; S. Rupprecht | 2022 | Neural distinctiveness of fatigue and low sleep quality in multiple sclerosis | Final Inclusion |
| A. Lazzarotto; M. Margoni; S. Franciotta; S. Zywicki; A. Riccardi; D. Poggiali; M. Anglani; P. Gallo | 2020 | Selective Cerebellar Atrophy Associates with Depression and Fatigue in the Early Phases of Relapse-Onset Multiple Sclerosis | Final Inclusion |
| A. M. Beaudoin; F. Rheault; G. Theaud; F. Laberge; K. Whittingstall; A. Lamontagne; M. Descoteaux | 2021 | Modern Technology in Multi-Shell Diffusion MRI Reveals Diffuse White Matter Changes in Young Adults With Relapsing-Remitting Multiple Sclerosis | Final Inclusion |
| A. Papadopoulou; N. Müller-Lenke; Y. Naegelin; G. Kalt; K. Bendfeldt; P. Kuster; M. Stoecklin; A. Gass; T. Sprenger; E. W. Radue; L. Kappos; I. K. Penner | 2013 | Contribution of cortical and white matter lesions to cognitive impairment in multiple sclerosis | Mixed MS: Do not report actual data, only stats values for RRMS group |
| A. Pokryszko-Dragan; A. Banaszek; M. Nowakowska-Kotas; K. Jeżowska-Jurczyk; E. Dziadkowiak; E. Gruszka; M. Zagrajek; M. Bilińska; S. Budrewicz; M. Sąsiadek; J. Bladowska | 2018 | Diffusion tensor imaging findings in the multiple sclerosis patients and their relationships to various aspects of disability | Final Inclusion |
| A. Romanello; S. Krohn; N. von Schwanenflug; C. Chien; J. Bellmann-Strobl; K. Ruprecht; F. Paul; C. Finke | 2022 | Functional connectivity dynamics reflect disability and multi-domain clinical impairment in patients with relapsing-remitting multiple sclerosis | Final Inclusion |
| A. Saberi; A. Abdolalizadeh; E. Mohammadi; M. A. Nahayati; H. Bagheri; B. Shekarchi; J. Kargar | 2021 | Thalamic shape abnormalities in patients with multiple sclerosis-related fatigue | Final Inclusion |
| A. Salter; K. Kowalec; K. C. Fitzgerald; G. Cutter; R. A. Marrie | 2020 | Comorbidity is associated with disease activity in MS: Findings from the CombiRx trial | Protocol paper |
| Akmaz, O., Koskderelioglu, A., Eskut, N., Sahan, B., Kusbeci, T. | 2022 | Restless legs syndrome in multiple sclerosis is related to retinal thinning | do not correlate MRI to D/F |
| Arm, J., Al-iedani, O., Ribbons, K., Lea, R., Lechner-Scott, J., Ramadan, S. | 2021 | Biochemical Correlations with Fatigue in Multiple Sclerosis Detected by MR 2D Localized Correlated Spectroscopy | no MRI |
| B. A. Parmenter; J. L. Shucard; D. W. Shucard | 2007 | Information processing deficits in multiple sclerosis: A matter of complexity | do not correlate MRI to D/F |

| | | | |
|---|---|---|---|
| B. A. Parmenter; R. Zivadinov; L. Kerenyi; R. Gavett; B. Weinstock-Guttman; M. G. Dwyer; N. Garg; F. Munschauer; R. H. B. Benedict | 2007 | Validity of the Wisconsin card sorting and Delis-Kaplan Executive Function System (DKEFS) sorting tests in multiple sclerosis | no D/F |
| B. Kis; B. Rumberg; P. Berlit | 2008 | Clinical characteristics of patients with late-onset multiple sclerosis | Mixed MS: Three different MS types but data is not differentiated |
| B. Nourbakhsh; C. Azevedo; J. Nunan-Saah; A. H. Maghzi; R. Spain; D. Pelletier; E. Waubant | 2016 | Longitudinal associations between brain structural changes and fatigue in early MS | Mixed MS: |
| Barešić, M., Reihl Crnogaj, M., Zadro, I., Anić, B. | 2021 | Demyelinating disease (multiple sclerosis) in a patient with psoriatic arthritis treated with adalimumab: a case-based review | Treatment: DMT |
| Benedict, R.H.B., Pol, J., Yasin, F., Hojnacki, D., Kolb, C., Eckert, S., Tacca, B., Drake, A., Wojcik, C., Morrow, S.A., Jakimovski, D., Fuchs, T.A., Dwyer, M.G., Zivadinov, R., Weinstock-Guttman, B. | 2021 | Recovery of cognitive function after relapse in multiple sclerosis | Treatment: Gel treatment/no fatigue or depression |
| C. Bauer; T. B. Dyrby; F. Sellebjerg; K. S. Madsen; O. Svolgaard; M. Blinkenberg; H. R. Siebner; K. W. Andersen | 2020 | Motor fatigue is associated with asymmetric connectivity properties of the corticospinal tract in multiple sclerosis | Final Inclusion |
| C. E. Schwartz; B. R. Quaranto; B. C. Healy; R. H. Benedict; T. L. Vollmer | 2013 | Cognitive reserve and symptom experience in multiple sclerosis: a buffer to disability progression over time? | wrong D/F test |
| C. Fazekas; M. Khalil; C. Enzinger; F. Matzer; S. Fuchs; F. Fazekas | 2013 | No impact of adult attachment and temperament on clinical variability in patients with clinically isolated syndrome and early multiple sclerosis | do not correlate MRI to D/F |
| C. Finke; J. Schlichting; S. Papazoglou; M. Scheel; A. Freing; C. Soemmer; L. M. Pech; A. Pajkert; C. Pfüller; J. T. Wuerfel; C. J. Ploner; F. Paul; A. U. Brandt | 2015 | Altered basal ganglia functional connectivity in multiple sclerosis patients with fatigue | Final Inclusion |
| C. J. Archibald; X. C. Wei; J. N. Scott; C. J. Wallace; Y. Zhang; L. M. Metz; J. R. Mitchell | 2004 | Posterior fossa lesion volume and slowed information processing in multiple sclerosis | Sample size <20 |
| C. Lebrun; C. Bensa; M. Debouverie; J. De Seze; S. Wiertlievski; B. Brochet; P. Clavelou; D. Brassat; P. Labauge; E. Roullet | 2008 | Unexpected multiple sclerosis: Follow-up of 30 patients with magnetic resonance imaging and clinical conversion profile | do not correlate MRI to D/F |
| C. Lebrun; O. H. Kantarci; A. Siva; D. Pelletier; D. T. Okuda | 2018 | Anomalies Characteristic of Central Nervous System Demyelination: Radiologically Isolated Syndrome | do not correlate MRI to D/F |
| Carandini, T., Mancini, M., Bogdan, I., Rae, C.L., Barritt, A.W., Clerico, M., Sethi, A., Harrison, N., Rashid, W., Scarpini, E., Galimberti, D., Bozzali, M., Cercignani, M. | 2021 | In vivo evidence of functional disconnection between brainstem monoaminergic nuclei and brain networks in multiple sclerosis | no D/F |
| Carotenuto, A., Valsasina, P., Preziosa, P., Mistri, D., Filippi, M., Rocca, M.A. | 2022 | Monoaminergic network abnormalities: A marker for multiple sclerosis-related fatigue and depression | Mixed MS: Did not separate |

| | | | |
|---|---|---|---|
| | | | between MS subgroups in analysis |
| Chylińska, M., Karaszewski, B., Komendziński, J., Wyszomirski, A., Sabisz, A., Halas, M., Szurowska, E. | 2022 | Skeletonized mean diffusivity and neuropsychological performance in relapsing-remitting multiple sclerosis | Treatment: DMT |
| D. A. Woo; M. J. Olek; E. M. Frohman | 2006 | Diagnosis and Management of Multiple Sclerosis: Case Studies | case studies |
| D. Iancheva; A. G. Trenova; K. Terziyski; S. Kandilarova; S. Mantarova | 2018 | Translational validity of PASAT and the effect of fatigue and mood in patients with relapsing remitting MS: A functional MRI study | Sample size <20 |
| D. Iancheva; A. Trenova; S. Mantarovau; K. Terziyski | 2019 | Functional Magnetic Resonance Imaging Correlations Between Fatigue and Cognitive Performance in Patients With Relapsing Remitting Multiple Sclerosis | Final Inclusion |
| Deverdun, J., Coget, A., Ayrignac, X., Carra-Dalliere, C., Krainik, A., Metzger, A., Labauge, P., Menjot de Champfleur, N., Le Bars, E. | 2021 | Cerebral Vasoreactivity as an Indirect MRI Marker of White Matter Tracts Alterations in Multiple Sclerosis | no D/F |
| E. M. Khedr; T. Desoky; A. Gamea; M. Y. Ezzeldin; A. F. Zaki | 2022 | Fatigue and brain atrophy in Egyptian patients with relapsing remitting multiple sclerosis | Final Inclusion |
| E. Portaccio; B. Goretti; V. Zipoli; B. Nacmias; M. L. Stromillo; M. L. Bartolozzi; G. Siracusa; L. Guidi; A. Federico; S. Sorbi; N. De Stefano; M. P. Amato | 2009 | APOE-epsilon 4 is not associated with cognitive impairment in relapsing-remitting multiple sclerosis | no D/F |
| E. Pravatà; C. Zecca; C. Sestieri; M. Caulo; G. C. Riccitelli; M. A. Rocca; M. Filippi; A. Cianfoni; C. Gobbi | 2016 | Hyperconnectivity of the dorsolateral prefrontal cortex following mental effort in multiple sclerosis patients with cognitive fatigue | Final Inclusion |
| E. Szabadi | 2013 | Functional neuroanatomy of the central noradrenergic system | no MRI |
| F. Al-Hussain; N. Al-Salloum; N. Alazwary; J. Saeedi; S. Howaidi; A. Daif | 2017 | Depression, anxiety and stress severities in multiple sclerosis patients using injectable versus oral treatments | do not correlate MRI to D/F |
| F. B. Tijhuis; T. A. A. Broeders; F. A. N. Santos; M. M. Schoonheim; J. Killestein; C. E. Leurs; Q. van Geest; M. D. Steenwijk; J. J. G. Geurts; H. E. Hulst; L. Douw | 2021 | Dynamic functional connectivity as a neural correlate of fatigue in multiple sclerosis | Final Inclusion |
| F. Morgante; V. Dattola; D. Crupi; M. Russo; V. Rizzo; M. F. Ghilardi; C. Terranova; P. Girlanda; A. Quartarone | 2011 | Is central fatigue in multiple sclerosis a disorder of movement preparation? | Final Inclusion |
| F. Patti; M. P. Amato; M. Trojano; S. Bastianello; M. R. Tola; B. Goretti; L. Caniatti; E. Di Monte; P. Ferrazza; V. B. Morra; S. Lo Fermo; O. Picconi; G. Luccichenti; C. S. Grp | 2009 | Cognitive impairment and its relation with disease measures in mildly disabled patients with relapsing-remitting multiple sclerosis: baseline results from the Cognitive Impairment in Multiple Sclerosis (COGIMUS) study | no D/F |
| F. Yousuf; G. Kim; S. Tauhid; B. I. Glanz; R. Chu; S. Tummala; B. C. Healy; R. Bakshi | 2016 | The contribution of cortical lesions to a composite MRI scale of disease severity in multiple sclerosis | do not correlate MRI to D/F |
| F. Zellini; G. Niepel; C. R. Tench; C. S. Constantinescu | 2009 | Hypothalamic involvement assessed by T1 relaxation time in patients with relapsing-remitting multiple sclerosis | Final Inclusion |

| | | | |
|---|---|---|---|
| F. Zhou; H. Gong; Q. Chen; B. Wang; Y. Peng; Y. Zhuang; C. S. Zee | 2016 | Intrinsic Functional Plasticity of the Thalamocortical System in Minimally Disabled Patients with Relapsing-Remitting Multiple Sclerosis | Final Inclusion |
| F. Zhou; Y. Zhuang; H. Gong; B. Wang; X. Wang; Q. Chen; L. Wu; H. Wan | 2014 | Altered inter-subregion connectivity of the default mode network in relapsing remitting multiple sclerosis: A functional and structural connectivity study | Final Inclusion |
| Fleischer, M., Schuh, H., Bickmann, N.M., Hagenacker, T., Krüger, K., Skripuletz, T., Fiedler, M., Kleinschnitz, C., Pul, R., Skuljec, J. | 2022 | Anti-EBNA1 IgG titre is not associated with fatigue in multiple sclerosis patients | no MRI |
| G. Bonnier; A. Roche; D. Romascano; S. Simioni; D. E. Meskaldji; D. Rotzinger; Y. C. Lin; G. Menegaz; M. Schluep; R. Du Pasquier; T. J. Sumpf; J. Frahm; J. P. Thiran; G. Krueger; C. Granziera | 2015 | Multicontrast MRI quantification of focal inflammation and degeneration in multiple sclerosis | do not correlate MRI to D/F |
| G. Niepel; R. Tench Ch; P. S. Morgan; N. Evangelou; D. P. Auer; C. S. Constantinescu | 2006 | Deep gray matter and fatigue in MS: a T1 relaxation time study | Final Inclusion |
| G. O. Nygaard; K. B. Walhovd; P. Sowa; J. L. Chepkoech; A. Bjørnerud; P. Due-Tønnessen; N. I. Landrø; S. Damangir; G. Spulber; A. B. Storsve; M. K. Beyer; A. M. Fjell; E. G. Celius; H. F. Harbo | 2015 | Cortical thickness and surface area relate to specific symptoms in early relapsing-remitting multiple sclerosis | Final Inclusion |
| G. Santangelo; M. D. Corte; M. Sparaco; G. Miele; F. Garramone; M. Cropano; S. Esposito; L. Lavorgna; A. Gallo; G. Tedeschi; S. Bonavita | 2021 | Coping strategies in relapsing-remitting multiple sclerosis non-depressed patients and their associations with disease activity | no D/F |
| G. Zito; E. Luders; L. Tomasevic; D. Lupoi; A. W. Toga; P. M. Thompson; P. M. Rossini; M. M. Filippi; F. Tecchio | 2014 | INTER-HEMISPHERIC FUNCTIONAL CONNECTIVITY CHANGES WITH CORPUS CALLOSUM MORPHOLOGY IN MULTIPLE SCLEROSIS | do not correlate MRI to D/F |
| Gilio, L., Fresegna, D., Gentile, A., Guadalupi, L., Sanna, K., De Vito, F., Balletta, S., Caioli, S., Rizzo, F.R., Musella, A., Iezzi, E., Moscatelli, A., Galifi, G., Fantozzi, R., Bellantonio, P., Furlan, R., Finardi, A., Vanni, V., Dolcetti, E., Bruno, A., Buttari, F., Mandolesi, G., Centonze, D., Stampanoni Bassi, M. | 2022 | Preventive exercise attenuates IL-2-driven mood disorders in multiple sclerosis | no MRI |
| Glasner P, Sabisz A, Chylińska M, Komendziński J, Wyszomirski A, Karaszewski B. | 2022 | Retinal nerve fiber and ganglion cell complex layer thicknesses mirror brain atrophy in patients with relapsing-remitting multiple sclerosis | do not correlate MRI to D/F |
| H. D. Keklikoğlu; T. K. Yoldaş; O. Zengin; E. B. Solak; S. Keskin | 2010 | Cognitive impairment in patients with early relapsing-remitting multiple sclerosis | no MRI |
| H. Hildebrandt; H. K. Hahn; J. A. Kraus; A. Schulte-Herbrüggen; B. Schwarze; G. Schwendemann | 2006 | Memory performance in multiple sclerosis patients correlates with central brain atrophy | Final Inclusion |
| H. Hildebrandt; P. Eling | 2014 | A longitudinal study on fatigue, depression, and their relation to neurocognition in multiple sclerosis | Final Inclusion |
| H. Joly; N. Capet; L. Mondot; M. Cohen; C. Suply; S. Bresch; C. Lebrun-Frenay | 2020 | Thalamic atrophy correlates with dysfunctional impulsivity in multiple sclerosis | do not correlate MRI to D/F |

| | | | |
|---|---|---|---|
| Healy, B.C., Glanz, B.I., Swallow, E., Signorovitch, J., Hagan, K., Silva, D., Pelletier, C., Chitnis, T., Weiner, H. | 2021 | Confirmed disability progression provides limited predictive information regarding future disease progression in multiple sclerosis | do not correlate MRI to D/F |
| Høgestøl, E.A., Ghezzo, S., Nygaard, G.O., Espeseth, T., Sowa, P., Beyer, M.K., Harbo, H.F., Westlye, L.T., Hulst, H.E., Alnæs, D. | 2022 | Functional connectivity in multiple sclerosis modelled as connectome stability: A 5-year follow-up study | do not correlate MRI to D/F |
| I. Håkansson; L. Johansson; C. Dahle; M. Vrethem; J. Ernerudh | 2019 | Fatigue scores correlate with other self-assessment data, but not with clinical and biomarker parameters, in CIS and RRMS | Mixed MS: CI and RRMS not separated for analysis |
| I. Specogna; F. Casagrande; A. Lorusso; M. Catalan; A. Gorian; L. Zugna; R. Longo; M. Zorzon; M. Naccarato; G. Pizzolato; M. Ukmar; M. A. Cova | 2012 | Functional MRI during the execution of a motor task in patients with multiple sclerosis and fatigue | Final Inclusion |
| J. I. Rojas; F. Sanchez; L. Patrucco; J. Miguez; C. Besada; E. Cristiano | 2017 | Brain structural changes in patients in the early stages of multiple sclerosis with depression | Final Inclusion |
| J. R. Abbatemarco; D. Ontaneda; K. Nakamura; S. Husak; Z. N. Wang; E. Alshehri; R. A. Bermel; D. S. Conway | 2020 | Comorbidity effect on processing speed test and MRI measures in multiple sclerosis patients | Mixed MS: MS not classified |
| J. Sepulcre; J. C. Masdeu; J. Goñi; G. Arrondo; N. Vélez de Mendizábal; B. Bejarano; P. Villoslada | 2009 | Fatigue in multiple sclerosis is associated with the disruption of frontal and parietal pathways | Mixed MS: MS not classified |
| J. Wilting; H. O. Rolfsnes; H. Zimmermann; M. Behrens; V. Fleischer; F. Zipp; A. Gröger | 2016 | Structural correlates for fatigue in early relapsing remitting multiple sclerosis | Final Inclusion |
| K. C. Kern; S. M. Gold; B. Lee; M. Montag; J. Horsfall; M. F. O'Connor; N. L. Sicotte | 2015 | Thalamic-hippocampal-prefrontal disruption in relapsing-remitting multiple sclerosis | no D/F |
| K. Konstantopoulos; M. Vikelis; J. A. Seikel; D. D. Mitsikostas | 2010 | The existence of phonatory instability in multiple sclerosis: An acoustic and electroglottographic study | do not correlate MRI to D/F |
| K. Makowiecki, , Stevens, N., Cullen, C.L., Zarghami, A., Nguyen, P.T., Johnson, L., Rodger, J., Hinder, M.R., Barnett, M., Young, K.M., Taylor, B.V. | 2022 | Safety of low-intensity repetitive transcranial magneTic brAin stimUlation foR people living with mUltiple Sclerosis (TAURUS): study protocol for a randomised controlled trial | Protocol paper |
| K. Okada; S. Kakeda; M. Tahara | 2020 | Olfactory identification associates with cognitive function and the third ventricle width in patients with relapsing-remitting multiple sclerosis | no D/F |
| K. Pierzchala; M. Adamczyk-Sowa; P. Dobrakowski; K. Kubicka-Baczyk; N. Niedziela; P. Sowa | 2015 | Demographic characteristics of MS patients in Poland's upper Silesia region | do not correlate MRI to D/F |
| K. Yarraguntla; F. Bao; S. Lichtman-Mikol; S. Razmjou; C. Santiago-Martinez; N. Seraji-Bozorgzad; S. Sriwastava; E. Bernitsas | 2019 | Characterizing Fatigue-Related White Matter Changes in MS: A Proton Magnetic Resonance Spectroscopy Study | Final Inclusion |
| K. Yarraguntla; N. Seraji-Bozorgzad; S. Lichtman-Mikol; S. Razmjou; F. Bao; S. Sriwastava; C. Santiago-Martinez; O. Khan; E. Bernitsas | 2018 | Multiple Sclerosis Fatigue: A Longitudinal Structural MRI and Diffusion Tensor Imaging Study | Final Inclusion |

| | | | |
|---|---|---|---|
| Kantorová, E., Hnilicová, P., Bogner, W., Grendár, M., Grossmann, J., Kováčová, S., Hečková, E., Strasser, B., Čierny, D., Zeleňák, K., Kurča, E. | 2022 | Neurocognitive performance in relapsing-remitting multiple sclerosis patients is associated with metabolic abnormalities of the thalamus but not the hippocampus– GABA-edited 1H MRS study | no MRI |
| Khedr, E.M., Abo-Elfetoh, N., Deaf, E., Hassan, H.M., Amin, M.T., Soliman, R.K., Attia, A.A., Zarzour, A.A., Zain, M., Mohamed-Hussein, A., Hashem, M.K., Hassany, S.M., Aly, A., Shoyb, A., Saber, M. | 2021 | Surveillance study of acute neurological manifestations among 439 egyptian patients with COVID-19 in assiut and Aswan University Hospitals | do not correlate MRI to D/F |
| Kopchak, O.O., Odintsova, T.A., Pulyk, O.R. | 2021 | COGNITIVE FUNCTIONS IN MULTIPLE SCLEROSIS PATIENTS DEPENDING ON THE DIFFERENT RISK FACTORS PRESENCE | can't access/non-English |
| Koubiyr, I., Dulau-Metras, C., Deloire, M., Charré-Morin, J., Saubusse, A., Brochet, B., Ruet, A. | 2022 | Amygdala network reorganization mediates the theory of mind performances in multiple sclerosis | do not correlate MRI to D/F |
| L. De Meijer; D. Merlo; O. Skibina; E. J. Grobbee; J. Gale; J. Haartsen; P. Maruff; D. Darby; H. Butzkueven; A. Van der Walt | 2018 | Monitoring cognitive change in multiple sclerosis using a computerized cognitive battery | Mixed MS: Do not distinguish between MS types |
| L. Debernard; T. R. Melzer; S. Alla; J. Eagle; S. Van Stockum; C. Graham; J. R. Osborne; J. C. Dalrymple-Alford; D. H. Miller; D. F. Mason | 2015 | Deep grey matter MRI abnormalities and cognitive function in relapsing-remitting multiple sclerosis | no D/F |
| L. Gilio; F. Buttari; L. Pavone; E. Iezzi; G. Galifi; E. Dolcetti; F. Azzolini; A. Bruno; A. Borrelli; M. Storto; R. Furlan; A. Finardi; T. Pekmezovic; J. Drulovic; G. Mandolesi; D. Fresegna; V. Vanni; D. Centonze; M. S. Bassi | 2022 | Fatigue in Multiple Sclerosis Is Associated with Reduced Expression of Interleukin-10 and Worse Prospective Disease Activity | Final Inclusion |
| L. Hofstetter; Y. Naegelin; L. Filli; P. Kuster; S. Traud; R. Smieskova; N. Mueller-Lenke; L. Kappos; A. Gass; T. Sprenger; I. K. Penner; T. E. Nichols; H. Vrenken; F. Barkhof; C. Polman; E. W. Radue; S. J. Borgwardt; K. Bendfeldt | 2014 | Progression in disability and regional grey matter atrophy in relapsing-remitting multiple sclerosis | wrong D/F test |
| L. Locatelli; R. Zivadinov; A. Grop; M. Zorzon | 2004 | Frontal parenchymal atrophy measures in multiple sclerosis | do not correlate MRI to D/F |
| L. Passamonti; A. Cerasa; M. Liguori; M. C. Gioia; P. Valentino; R. Nistico; A. Quattrone; F. Fera | 2009 | Neurobiological mechanisms underlying emotional processing in relapsing-remitting multiple sclerosis | Sample size <20 |
| L. Tomasevic; G. Zito; P. Pasqualetti; M. Filippi; D. Landi; A. Ghazaryan; D. Lupoi; C. Porcaro; F. Bagnato; P. Rossini; F. Tecchio | 2013 | Cortico-muscular coherence as an index of fatigue in multiple sclerosis | Final Inclusion |
| L. Wu; M. Huang; F. Zhou; X. Zeng; H. Gong | 2020 | Distributed causality in resting-state network connectivity in the acute and remitting phases of RRMS | Final Inclusion |
| L. Wu; Y. Zhang; F. Q. Zhou; L. Gao; L. C. He; X. J. Zeng; H. H. Gong | 2016 | Altered intra- and interregional synchronization in relapsing-remitting multiple sclerosis: a resting-state fMRI study | Final Inclusion |

| | | | |
|---|---|---|---|
| Labbe, T.P., Montalba, C., Zurita, M., Ciampi, E.L., Cruz, J.P., Vasquez, M., Uribe, S., Crossley, N., Cárcamo, C. | 2021 | Regional brain atrophy is related to social cognition impairment in multiple sclerosis [La atrofia cerebral regional se relaciona con el deterioro de la cognición social en sclerosis múltiple] | do not correlate MRI to D/F |
| M. A. Rocca; A. Meani; G. C. Riccitelli; B. Colombo; M. Rodegher; A. Falini; G. Comi; M. Filippi | 2016 | Abnormal adaptation over time of motor network recruitment in multiple sclerosis patients with fatigue | Final Inclusion |
| M. A. Rocca; M. Absinta; P. Valsasina; M. Copetti; D. Caputo; G. Comi; M. Filippi | 2012 | Abnormal cervical cord function contributes to fatigue in multiple sclerosis | Spinal cord |
| M. A. Rocca; R. Gatti; F. Agosta; P. Broglia; P. Rossi; E. Riboldi; M. Corti; G. Comi; M. Filippi | 2009 | Influence of task complexity during coordinated hand and foot movements in MS patients with and without fatigue. A kinematic and functional MRI study | Final Inclusion |
| M. A. Wojtowicz; Y. Ishigami; E. L. Mazerolle; J. D. Fisk | 2014 | Stability of intraindividual variability as a marker of neurologic dysfunction in relapsing remitting multiple sclerosis | Sample size <20 |
| M. C. Bonnet; M. S. A. Deloire; E. Salort; V. Dousset; K. G. Petry; B. Brochet | 2006 | Evidence of cognitive compensation associated with educational level in early relapsing-remitting multiple sclerosis | do not correlate MRI to D/F |
| M. Calabrese; F. Rinaldi; P. Grossi; I. Mattisi; V. Bernardi; A. Favaretto; P. Perini; P. Gallo | 2010 | Basal ganglia and frontal/parietal cortical atrophy is associated with fatigue in relapsing-remitting multiple sclerosis | Final Inclusion |
| M. Cavallari; M. Palotai; B. I. Glanz; S. Egorova; J. C. Prieto; B. C. Healy; T. Chitnis; C. R. G. Guttmann | 2016 | Fatigue predicts disease worsening in relapsing-remitting multiple sclerosis patients | Final Inclusion |
| M. Codella; M. A. Rocca; B. Colombo; F. Martinelli-Boneschi; G. Comi; M. Filippi | 2002 | Cerebral grey matter pathology and fatigue in patients with multiple sclerosis: a preliminary study | Final Inclusion |
| M. Filippi; M. A. Rocca; B. Colombo; A. Falini; M. Codella; G. Scotti; G. Comi | 2002 | Functional magnetic resonance imaging correlates of fatigue in multiple sclerosis | Final Inclusion |
| M. Gschwind; M. Hardmeier; D. Van De Ville; M. I. Tomescu; I. K. Penner; Y. Naegelin; P. Fuhr; C. M. Michel; M. Seeck | 2016 | Fluctuations of spontaneous EEG topographies predict disease state in relapsing-remitting multiple sclerosis | do not correlate MRI to D/F |
| M. Huang; F. Zhou; L. Wu; B. Wang; H. Wan; F. Li; X. Zeng; H. Gong | 2018 | Synchronization within, and interactions between, the default mode and dorsal attention networks in relapsing-remitting multiple sclerosis | Final Inclusion |
| M. Inglese; S. J. Park; G. Johnson; J. S. Babb; L. Miles; H. Jaggi; J. Herbert; R. I. Grossman | 2007 | Deep gray matter perfusion in multiple sclerosis: Dynamic susceptibility contrast perfusion magnetic resonance imaging at 3 T | wrong D/F test |
| M. J. Fartaria; K. O'Brien; A. Sorega; G. Bonnier; A. Roche; P. Falkovskiy; G. Krueger; T. Kober; M. B. Cuadra; C. Granziera | 2017 | An Ultra-High Field Study of Cerebellar Pathology in Early Relapsing-Remitting Multiple Sclerosis Using MP2RAGE | no D/F |
| M. Jehna; C. Langkammer; M. Wallner-Blazek; C. Neuper; M. Loitfelder; S. Ropele; S. Fuchs; M. Khalil; A. Pluta-Fuerst; F. Fazekas; C. Enzinger | 2011 | Cognitively preserved MS patients demonstrate functional differences in processing neutral and emotional faces | do not correlate MRI to D/F |
| M. L. Polliack; Y. Barak; A. Achiron | 2001 | Late-onset multiple sclerosis | no MRI |

| | | | |
|---|---|---|---|
| M. N. Burns; E. Nawacki; M. J. Kwasny; D. Pelletier; D. C. Mohr | 2014 | Do positive or negative stressful events predict the development of new brain lesions in people with multiple sclerosis? | no D/F |
| M. P. Amato; E. Portaccio; B. Goretti; V. Zipoli; A. Iudice; D. D. Pina; G. Malentacchi; S. Sabatini; P. Annunziata; M. Falcini; M. Mazzoni; M. Mortilla; C. Fonda; N. De Stefano | 2010 | Relevance of cognitive deterioration in early relapsing-remitting MS: A 3-year follow-up study | no D/F |
| M. Pardini; L. Bonzano; G. L. Mancardi; L. Roccatagliata | 2010 | Frontal networks play a role in fatigue perception in multiple sclerosis | Final Inclusion |
| M. Pardini; L. Bonzano; M. Bergamino; G. Bommarito; P. Feraco; A. Murugavel; M. Bove; G. Brichetto; A. Uccelli; G. Mancardi; L. Roccatagliata | 2015 | Cingulum bundle alterations underlie subjective fatigue in multiple sclerosis | Final Inclusion |
| M. Russo; A. Calamuneri; A. Cacciola; L. Bonanno; A. Naro; V. Dattola; E. Sessa; M. Buccafusca; D. Milardi; P. Bramanti; R. S. Calabro; G. Anastasi; A. Quartarone | 2017 | Neural correlates of fatigue in multiple sclerosis: a combined neurophysiological and neuroimaging approach (R1) | can't access |
| M. Stangel; I. K. Penner; B. A. Kallmann; C. Lukas; B. C. Kieseier | 2015 | Towards the implementation of ‘no evidence of disease activity' in multiple sclerosis treatment: The multiple sclerosis decision model | no D/F |
| M. Summers; J. Swanton; K. Fernando; C. Dalton; D. H. Miller; L. Cipolotti; M. A. Ron | 2008 | Cognitive impairment in multiple sclerosis can be predicted by imaging early in the disease | do not correlate MRI to D/F |
| M. Yildiz; F. Brugger; N. Kästle; B. Tettenborn | 2016 | Neurocognitive impairment is associated with corpus callosum atrophy in multiple sclerosis | Mixed MS: Includes other MS types in the MS group |
| M. Zorzon; R. Zivadinov; L. Locatelli; B. Stival; D. Nasuelli; A. Bratina; A. Bosco; M. A. Tommasi; R. S. Pozzi Mucelli; M. Ukmar; G. Cazzato | 2003 | Correlation of sexual dysfunction and brain magnetic resonance imaging in multiple sclerosis | do not correlate MRI to D/F |
| Manca, R., Mitolo, M., Wilkinson, I., Paling, D., Sharrack, B., Venneri, A. | 2021 | A network-based cognitive training induces cognitive improvements and neuroplastic changes in patients with relapsing-remitting multiple sclerosis: An exploratory case-control study | Treatmen: non-drug intervention study |
| N. Bergsland; R. Zivadinov; M. G. Dwyer; B. Weinstock-Guttman; R. H. B. Benedict | 2016 | Localized atrophy of the thalamus and slowed cognitive processing speed in MS patients | do not correlate MRI to D/F |
| N. Derache; B. Grassiot; F. Mézenge; A. Emmanuelle Dugué; B. Desgranges; J. M. Constans; G. L. Defer | 2013 | Fatigue is associated with metabolic and density alterations of cortical and deep gray matter in Relapsing-Remitting-Multiple Sclerosis patients at the earlier stage of the disease: A PET/MR study | wrong D/F test |
| N. Téllez; J. Alonso; J. Río; M. Tintoré; C. Nos; X. Montalban; A. Rovira | 2008 | The basal ganglia: a substrate for fatigue in multiple sclerosis | Final Inclusion |
| Nabizadeh, F., Balabandian, M., Rostami, M.R., Owji, M., Sahraian, M.A., Bidadian, M., Ghadiri, F., Rezaeimanesh, N., Moghadasi, A.N. | 2022 | Association of cognitive impairment and quality of life in patients with multiple sclerosis: A cross-sectional study | do not correlate MRI to D/F |

| | | | |
|---|---|---|---|
| Nath, S.R., Grewal, P., Cho, T., Mao-Draayer, Y. | 2022 | Familial multiple sclerosis in patients with Von Hippel-Lindau disease | do not correlate MRI to D/F |
| Newland, P., Chen, L., Sun, P., Zempel, J. | 2021 | Neurophysiological Correlates of Fatigue in Multiple Sclerosis | Sample size <20 |
| O. O. Kopchak; T. A. Odintsova | 2021 | Cognitive impairment and depression in patients with relapsing-remitting multiple sclerosis depending on age and neuroimaging findings | Final Inclusion |
| O. Svolgaard; K. W. Andersen; C. Bauer; K. H. Madsen; M. Blinkenberg; F. Selleberg; H. R. Siebner | 2018 | Cerebellar and premotor activity during a non-fatiguing grip task reflects motor fatigue in relapsing-remitting multiple sclerosis | Final Inclusion |
| O. Svolgaard; K. W. Andersen; C. Bauer; K. H. Madsen; M. Blinkenberg; F. Sellebjerg; H. R. Siebner | 2022 | Mapping grip-force related brain activity after a fatiguing motor task in multiple sclerosis | Final Inclusion |
| Ö. Yaldizli; I. K. Penner; T. Yonekawa; Y. Naegelin; J. Kuhle; M. Pardini; D. T. Chard; C. Stippich; J. I. Kira; K. Bendfeldt; M. Amann; E. W. Radue; L. Kappos; T. Sprenger | 2016 | The association between olfactory bulb volume, cognitive dysfunction, physical disability and depression in multiple sclerosis | Final Inclusion |
| Ö. Yaldizli; S. Glassl; D. Sturm; A. Papadopoulou; A. Gass; B. Tettenborn; N. Putzki | 2011 | Fatigue and progression of corpus callosum atrophy in multiple sclerosis | Final Inclusion |
| Ogisu, K., Niino, M., Miyazaki, Y., Kikuchi, S. | 2021 | Optimal indicator for histogram analysis of fractional anisotropy for normal-appearing white matter in multiple sclerosis | Sample size <20 |
| Ooi, S., Kalincik, T., Perucca, P., Monif, M. | 2021 | The prevalence of epileptic seizures in multiple sclerosis in a large tertiary hospital in Australia | do not correlate MRI to D/F |
| P. Puz; A. Lasek-Bal; A. Steposz; K. Bartoszek | 2018 | Effect of comorbidities on the course of multiple sclerosis | do not correlate MRI to D/F |
| Palotai, M., Wallack, M., Kujbus, G., Dalnoki, A., Guttmann, C. | 2021 | Usability of a mobile app for real-time assessment of fatigue and related symptoms in patients with multiple sclerosis: Observational study | no MRI |
| Parray, Z., Zargar, M.H., Asimi, R., Dar, W.R., Yaqoob, A., Raina, A., Ganie, H., Wani, M., Shah, Z.A. | 2022 | Interleukin 32 gene promoter polymorphism: A genetic risk factor for multiple sclerosis in Kashmiri population | no MRI |
| R. Riccelli; L. Passamonti; A. Cerasa; S. Nigro; S. M. Cavalli; C. Chiriaco; P. Valentino; R. Nisticò; A. Quattrone | 2016 | Individual differences in depression are associated with abnormal function of the limbic system in multiple sclerosis patients | Final Inclusion |
| R. Righart; V. Biberacher; L. E. Jonkman; R. Klaver; P. Schmidt; D. Buck; A. Berthele; J. S. Kirschke; C. Zimmer; B. Hemmer; J. J. G. Geurts; M. Mühlau | 2017 | Cortical pathology in multiple sclerosis detected by the T1/T2-weighted ratio from routine magnetic resonance imaging | do not correlate MRI to D/F |
| R. Zivadinov; J. Sepcic; D. Nasuelli; R. De Masi; L. M. Bragadin; M. A. Tommasi; S. Zambito-Marsala; R. Moretti; A. Bratina; M. Ukmar; R. S. Pozzi-Mucelli; A. Grop; G. Cazzato; M. Zorzon | 2001 | A longitudinal study of brain atrophy and cognitive disturbances in the early phase of relapsing-remitting multiple sclerosis | do not correlate MRI to D/F |
| R. Zivadinov; M. Zorzon; L. Locatelli; B. Stival; F. Monti; D. Nasuelli; M. A. Tommasi; A. Bratina; G. Cazzato | 2003 | Sexual dysfunction in multiple sclerosis: A MRI, neurophysiological and urodynamic study | no D/F |

| | | | |
|---|---|---|---|
| Rocca, M.A., Valsasina, P., Colombo, B., Martinelli, V., Filippi, M. | 2021 | Cortico-subcortical functional connectivity modifications in fatigued multiple sclerosis patients treated with fampridine and amantadine | Treatment: DMT |
| Rojas, J.I., Patrucco, L., Alonso, R., Garcea, O., Deri, N., Carnero Contentti, E., Lopez, P.A., Pettinicchi, J.P., Caride, A., Cristiano, E. | 2021 | Diagnostic uncertainty during the transition to secondary progressive multiple sclerosis: Multicenter study in Argentina | do not correlate MRI to D/F |
| S. A. Mohamed; O. El-Deib | 2014 | Depressive symptoms as a predictor of outcome in patients with multiple sclerosis | no MRI |
| S. Barone; M. E. Caligiuri; P. Valentino; A. Cherubini; C. Chiriaco; A. Granata; E. Filippelli; T. Tallarico; R. Nistico; A. Quattrone | 2018 | Multimodal assessment of normal-appearing corpus callosum is a useful marker of disability in relapsing-remitting multiple sclerosis: an MRI cluster analysis study | do not correlate MRI to D/F |
| S. Cader; A. Cifelli; Y. Abu-Omar; J. Palace; P. M. Matthews | 2006 | Reduced brain functional reserve and altered functional connectivity in patients with multiple sclerosis | no D/F |
| S. Collorone; N. Cawley; F. Grussu; F. Prados; F. Tona; A. Calvi; B. Kanber; T. Schneider; L. Kipp; H. Zhang; D. C. Alexander; A. J. Thompson; A. Toosy; C. A. M. G. Wheeler-Kingshott; O. Ciccarelli | 2020 | Reduced neurite density in the brain and cervical spinal cord in relapsing–remitting multiple sclerosis: A NODDI study | no D/F |
| S. Golde; J. Heine; J. Pöttgen; M. Mantwill; S. Lau; K. Wingenfeld; C. Otte; I. K. Penner; A. K. Engel; C. Heesen; J. P. Stellmann; I. Dziobek; C. Finke; S. M. Gold | 2020 | Distinct Functional Connectivity Signatures of Impaired Social Cognition in Multiple Sclerosis | Final Inclusion |
| S. Jaeger; F. Paul; M. Scheel; A. Brandt; J. Heine; D. Pach; C. M. Witt; J. Bellmann-Strobl; C. Finke | 2019 | Multiple sclerosis-related fatigue: Altered resting-state functional connectivity of the ventral striatum and dorsolateral prefrontal cortex | Final Inclusion |
| S. M. Gold; K. C. Kern; M. F. O'Connor; M. J. Montag; A. Kim; Y. S. Yoo; B. S. Giesser; N. L. Sicotte | 2010 | Smaller cornu ammonis 2-3/dentate gyrus volumes and elevated cortisol in multiple sclerosis patients with depressive symptoms | Final Inclusion |
| S. Nigro; L. Passamonti; R. Riccelli; N. Toschi; F. Rocca; P. Valentino; R. Nisticò; F. Fera; A. Quattrone | 2015 | Structural 'connectomic' alterations in the limbic system of multiple sclerosis patients with major depression | Final Inclusion |
| S. P. Hojjat; C. G. Cantrell; T. J. Carroll; R. Vitorino; A. Feinstein; L. Zhang; S. P. Symons; S. A. Morrow; L. Lee; P. O'Connor; R. I. Aviv | 2016 | Perfusion reduction in the absence of structural differences in cognitively impaired versus unimpaired RRMS patients | do not correlate MRI to D/F |
| S. Rossi; V. Studer; C. Motta; S. Polidoro; J. Perugini; G. Macchiarulo; A. M. Giovannetti; L. Pareja-Gutierrez; A. Calò; I. Colonna; R. Furlan; G. Martino; D. Centonze | 2017 | Neuroinflammation drives anxiety and depression in relapsing-remitting multiple sclerosis | do not correlate MRI to D/F |
| S. Sevim | 2016 | Relapses in multiple sclerosis: Definition, pathophysiology, features, imitators, and treatment | no D/F |
| Saruhan, E., Korkmaz, M., Altiparmak, B., Tosun, K., Kutlu, G. | 2022 | COMPARISON OF OREXIN-A AND NEUROFILAMENT LIGHT CHAIN LEVELS IN PATIENTS WITH RELAPSING-REMITTING MULTIPLE SCLEROSIS: A PILOT STUDY [OREXIN-A- ÉS NEUROFILAMENTUM- | no MRI |

| | | | |
|---|---|---|---|
| | | KÖNNYÛLÁNC FEHÉRJESZINTEK RELAPSZÁLÓ-REMITTÁLÓ SCLEROSIS MULTIPLEXBEN SZENVEDÔKNÉL: PILOT VIZSGÁLAT] | |
| Soares, J.M., Conde, R., Magalhães, R., Marques, P., Magalhães, R., Gomes, L., Gonçalves, Ó.F., Arantes, M., Sampaio, A. | 2021 | Alterations in functional connectivity are associated with white matter lesions and information processing efficiency in multiple sclerosis | do not correlate MRI to D/F |
| Stascheit, F., Li, L., Mai, K., Baum, K., Siebert, E., Ruprecht, K. | 2021 | Delayed onset hypophysitis after therapy with daclizumab for multiple sclerosis – A report of two cases | Treatment: DMT |
| T. A. Hassan; S. F. Elkholy; B. E. Mahmoud; M. ElSherbiny | 2019 | Multiple sclerosis and depressive manifestations: can diffusion tensor MR imaging help in the detection of microstructural white matter changes? | Final Inclusion |
| T. K. Yoldas; H. D. Keklikoglu; O. Zengin; E. B. Solak; S. Keskin | 2010 | Relation of Serum Uric Acid Level with Cognitive Functions and Number of Plaques in Patients with Relapsing-Remitting Multiple Sclerosis | no D/F |
| T. Štecková; P. Hluštík; V. Sládková; F. Odstrčil; J. Mareš; P. Kaňovský | 2014 | Thalamic atrophy and cognitive impairment in clinically isolated syndrome and multiple sclerosis | Final Inclusion |
| Trufanov, A., Bisaga, G., Skulyabin, D., Temniy, A., Poplyak, M., Chakchir, O., Efimtsev, A., Dmitriy, T., Odinak, M., Litvinenko, I. | 2021 | Thalamic nuclei degeneration in multiple sclerosis | do not correlate MRI to D/F |
| V. Biberacher; C. C. Boucard; P. Schmidt; C. Engl; D. Buck; A. Berthele; M. M. Hoshi; C. Zimmer; B. Hemmer; M. Muhlau | 2015 | Atrophy and structural variability of the upper cervical cord in early multiple sclerosis | Spinal cord |
| V. M. Leavitt; E. De Meo; G. Riccitelli; M. A. Rocca; G. Comi; M. Filippi; J. F. Sumowski | 2015 | Elevated body temperature is linked to fatigue in an Italian sample of relapsing-remitting multiple sclerosis patients | do not correlate MRI to D/F |
| V. Martinovic; I. Nikolic; S. Mesaros; J. Drulovic | 2020 | Bilateral horizontal gaze palsy in benign multiple sclerosis | no D/F |
| Waliszewska-Prosół, M., Nowakowska-Kotas, M., Misiak, B., Chojdak-Łukasiewicz, J., Budrewicz, S., Pokryszko-Dragan, A. | 2022 | Allostatic load index in patients with multiple sclerosis: A case-control study | no MRI |
| Y. Benesova; I. Niedermayerova; M. Mechl; P. Havlikova | 2003 | The relation between brain MRI lesions and depressive symptoms in multiple sclerosis | Final Inclusion |
| Y. Shen; L. Bai; Y. Gao; F. Cui; Z. Tan; Y. Tao; C. Sun; L. Zhou | 2014 | Depressive symptoms in multiple sclerosis from an in vivo study with TBSS | Sample size <20 |
| Yalachkov, Y., Anschuetz, V., Jakob, J., Schaller-Paule, M.A., Schaefer, J.H., Reilaender, A., Friedauer, L., Behrens, M., Foerch, C. | 2021 | C-Reactive Protein Levels and Gadolinium-Enhancing Lesions Are Associated With the Degree of Depressive Symptoms in Newly Diagnosed Multiple Sclerosis | Mixed MS: Did not separate between MS subgroups in analysis |
| Zanghì, A., D'Amico, E., Lo Fermo, S., Patti, F. | 2021 | Exploring polypharmacy phenomenon in newly diagnosed relapsing–remitting multiple sclerosis: a cohort ambispective single-centre study | Treatment: DMT |
