## Supplemental Text 1 for "Brain connectivity changes underlying depression and fatigue in relapsing-remitting multiple sclerosis: a systematic review"

### 1. Supplement Methods

#### 1.1 Quality assessment

The Institute of Health Economics (IHE) 'Quality Appraisal of Case Series Studies Checklist' was used to assess the quality of the longitudinal studies included as it can be used for longitudinal designs [1]. 'The checklist has seven categories: study objective, study design, study population, outcome measure, statistical analysis, results and conclusions, competing interests and sources of support [1] (S2 and S3 Tables). The checklist was modified by removing five questions that did not apply to included longitudinal studies. To aid the comparison of the quality of the included studies, we awarded a point for each 'Yes' answer, 0.5 point for 'Partial', and no points for each 'No' or 'Unclear' answer. No studies received full points (S2 and S4 Tables).

The 'Appraisal tool for cross sectional studies' (AXIS) was used to assess quality of cross-sectional studies [2]. The AXIS tool is a relatively new tool that was developed specifically for cross-sectional studies, as many pre-existing tools focus on randomised control trials or other longitudinal study designs [2]. The AXIS tool consists of 20 questions assessing all sections of the paper for clarity, sampling bias, data analysis, result presentation, interpretation of results and conflicts of interest. For this review, three questions regarding information about non-responders were excluded as they were not appropriate for the types of studies assessed. AXIS scores for each assessed study were summarised into sections, including 'study design', 'selection bias', 'comparability/methods', and 'outcomes' (see S3 Table for individual scores). A higher AXIS score indicates a higher quality study.

Two reviewers conducted the quality assessment independently. In cases of disagreement on scores, all papers were re-evaluated by both reviewers, and discussed to achieve consensus. Consensus scores were reported.

### 2. Supplement Results

#### 2.1 Depression assessment

##### 2.1.1 Conventional MRI measures

Most of studies (14/17) used the Beck Depression Index (BDI) to assess depression severity. The remaining studies used either Hamilton Depression Rating Scale (HDRS) [3, 4], Centre for Epidemiological Studies – Depression (CES-D) [5], or Hospital Anxiety and Depression Scale (HADS) [6]. Additionally, three papers used Diagnostic and Statistical Manual of Mental Disorders, 5th Edition (DSM-V) in addition to the BDI [7-9].

##### 2.1.2 Structural connectivity

One study used HADS-D [6], one used DSM-V [10], one – BDI [11], and two studies used the DSM-V and BDI together [8, 9].

##### 2.1.3 Functional connectivity

Two studies used BDI [12, 13], one used BDI with DSM-V [7], and the other two studies used HDRS [4] and HADS-D [6].

#### 2.2 Fatigue assessment

##### 2.2.1 Conventional MRI measures

23/46 used the Fatigue Severity Scale (FSS), 13/46 used the Modified Fatigue Impact Scale (MFIS), 6/46 studies used the Fatigue Scale for Motor and Cognitive Functions (FSMC) [6, 14], 3/46 used both FSS and MFIS, and one used CIS-20r (Table 5).

##### 2.2.2 Structural connectivity

Six out of fifteen studies used the FSS, 5/15 studies used MFIS and 3/13 used FSMC, and one - FIS (Table 5).

##### 2.2.3 Functional connectivity

Seven out of twenty studies used the FSS, 8/20 used the MFIS, one used CIS-20r and 4/20 used the FSMC (Table 5).

10. Hassan TA, Elkholy SF, Mahmoud BE, ElSherbiny M. Multiple sclerosis and depressive manifestations: can diffusion tensor MR imaging help in the detection of microstructural white

matter changes? Egyptian Journal of Radiology and Nuclear Medicine. 2019;50(1). doi: 10.1186/s43055-019-0033-8. PubMed PMID: WOS:000486165900001.
11. Beaudoin AM, Rheault F, Theaud G, Laberge F, Whittingstall K, Lamontagne A, et al. Modern Technology in Multi-Shell Diffusion MRI Reveals Diffuse White Matter Changes in Young Adults With Relapsing-Remitting Multiple Sclerosis. Frontiers in Neuroscience. 2021;15:13. doi: 10.3389/fnins.2021.665017. PubMed PMID: WOS:000687832800001.
12. Jaeger S, Paul F, Scheel M, Brandt A, Heine J, Pach D, et al. Multiple sclerosis-related fatigue: Altered resting-state functional connectivity of the ventral striatum and dorsolateral prefrontal cortex. Mult Scler. 2019;25(4):554-64. Epub 20180221. doi: 10.1177/1352458518758911. PubMed PMID: 29464981.
13. Romanello A, Krohn S, von Schwanenflug N, Chien C, Bellmann-Strobl J, Ruprecht K, et al. Functional connectivity dynamics reflect disability and multi-domain clinical impairment in patients with relapsing-remitting multiple sclerosis. Neuroimage Clin. 2022;36:103203. Epub 20220916. doi: 10.1016/j.nicl.2022.103203. PubMed PMID: 36179389; PubMed Central PMCID: PMCPMC9668632.
14. Bauer C, Dyrby TB, Sellebjerg F, Madsen KS, Svolgaard O, Blinkenberg M, et al. Motor fatigue is associated with asymmetric connectivity properties of the corticospinal tract in multiple sclerosis. Neuroimage-Clinical. 2020;28. doi: 10.1016/j.nicl.2020.102393. PubMed PMID: WOS:000600619100036.
