## Supplemental Table 2 for "Brain connectivity changes underlying depression and fatigue in relapsing-remitting multiple sclerosis: a systematic review"

**S2 Table. Data extraction table.**

| DOI | Paper | Author | Type of study - Longitudinal or cross sectional | Part of brain studied | MRI measures | MRI full report | Machine + Field strength | Participants - N for each group | Participants age range; Median/mean age | EDSS score median/mean (or range if median not given) | Fatigue - What assessment method was used and if cut off values were used | Depression - What assessment method was used: if cut off values were used | Major findings - Differences (or no differences) between groups and in what regions (p-values, CI, correlation coefficient, etc.) | Comments |
|---|---|---|---|---|---|---|---|---|---|---|---|---|---|---|
