## Supplemental Table 3 for "Brain connectivity changes underlying depression and fatigue in relapsing-remitting multiple sclerosis: a systematic review"

**S3 Table**. **Quality assessment of cross-sectional studies using the 'Appraisal tool of cross-sectional studies' (AXIS) [1].** Percentages are based on the number of 'positively' answered questions per category, with 17 questions in total. A high percentage indicates higher quality. A cut-off for percentage of 'positive' answers indicating a higher quality is not defined. AXIS merely allows for comparison between studies of similar design.

| **Author** | **Study design** | **Selection bias** | **Comparability / Method** | **Outcomes** | **Total Score** |
|---|---|---|---|---|---|
| Alshehri et al. [2] | 4 (100%) | 3 (75%) | 5 (100%) | 4 (100%) | 16 (94%) |
| Altermatt et al. [3] | 4 (100%) | 3 (75%) | 5 (100%) | 4 (100%) | 16 (94%) |
| Andreasen et al. [4] | 4 (100%) | 3 (75%) | 5 (100%) | 4 (100%) | 16 (94%) |
| Bauer et al. [5] | 3 (75%) | 3 (75%) | 5 (100%) | 4 (100%) | 15 (88%) |
| Beaudoin et al. [6] | 4 (100%) | 3 (75%) | 5 (100%) | 4 (100%) | 16 (94%) |
| Benesova et al. [7] | 2 (50%) | 3 (75%) | 4 (80%) | 3 (75%) | 12 (71%) |
| Bisecco et al. [8] | 4 (100%) | 3 (75%) | 5 (100%) | 4 (100%) | 16 (94%) |
| Carotenuto et al. [9] | 4 (100%) | 3 (75%) | 5 (100%) | 4 (100%) | 16 (94%) |
| Codella et al. [10] | 2 (50%) | 3 (75%) | 5 (100%) | 3 (75%) | 13 (76%) |
| Damasceno et al. [11] | 4 (100%) | 3 (75%) | 5 (100%) | 3 (75%) | 15 (88%) |
| Filippi et al. [12] | 3 (75%) | 3 (75%) | 5 (100%) | 4 (100%) | 15 (88%) |
| Finke et al. [13] | 3 (75%) | 3 (75%) | 5 (100%) | 4 (100%) | 15 (88%) |
| Gold et al. [14] | 4 (100%) | 3 (75%) | 5 (100%) | 4 (100%) | 16 (94%) |
| Golde et al. [15] | 4 (100%) | 3 (75%) | 5 (100%) | 4 (100%) | 16 (94%) |
| Cruz Gomez et al. [16] | 4 (100%) | 3 (75%) | 4 (80%) | 4 (100%) | 15 (88%) |
| Hassan et al. [17] | 4 (100%) | 3 (75%) | 3 (60%) | 4 (100%) | 13 (76%) |
| Hildebrandt et al. [18] | 4 (100%) | 3 (75%) | 5 (100%) | 3 (75%) | 15 (88%) |
| Huang et al. [19] | 4 (100%) | 3 (75%) | 5 (100%) | 4 (100%) | 16 (94%) |
| Iancheva et al. [20] | 4 (100%) | 3 (75%) | 4 (80%) | 3 (75%) | 14 (82%) |
| Jaeger et al. [21] | 4 (100%) | 3 (75%) | 5 (100%) | 4 (100%) | 16 (94%) |
| Kever et al. [22] | 4 (100%) | 3 (75%) | 4 (80%) | 4 (100%) | 15(88%) |
| Khedr et al. [23] | 4 (100%) | 3 (75%) | 5 (100%) | 4 (100%) | 16 (94%) |
| Kopchak and Odintsova [22] | 3 (75%) | 3 (75%) | 1 (20%) | 3 (75%) | 10 (59%) |
| Lazzarotto et al. [24] | 4 (100%) | 3 (75%) | 5 (100%) | 3 (75%) | 15 (88%) |
| Morgante et al. [25] | 4 (100%) | 3 (75%) | 5 (100%) | 4 (100%) | 16 (94%) |
| Niepel et al. [26] | 3 (75%) | 3 (75%) | 5 (100%) | 3 (75%) | 14 (82%) |
| Nigro et al. [27] | 4 (100%) | 3 (75%) | 5 (100%) | 4 (100%) | 16 (94%) |
| Nygaard et al. [28] | 4 (100%) | 3 (75%) | 5 (100%) | 4 (100%) | 16 (94%) |
| Pardini et al. [29] | 3 (75%) | 3 (75%) | 5 (100%) | 3 (75%) | 14 (82%) |
| Pardini et al. [30] | 3 (75%) | 3 (75%) | 3 (75%) | 4 (100%) | 13 (76%) |
| Pokryszko-Dragan et al. [31] | 4 (100%) | 3 (75%) | 5 (100%) | 4 (100%) | 16 (94%) |
| Pravatà et al. [32] | 4 (100%) | 3 (75%) | 5 (100%) | 4 (100%) | 16 (94%) |
| Riccelli et al. [33] | 4 (100%) | 3 (75%) | 5 (100%) | 4 (100%) | 16 (94%) |
| Rocca et al. [34] | 4 (100%) | 3 (75%) | 5 (100%) | 4 (100%) | 16 (94%) |
| Rocca et al. [35] | 4 (100%) | 3 (75%) | 5 (100%) | 4 (100%) | 16 (94%) |
| Rojas et al. [36] | 3 (75%) | 3 (75%) | 5 (100%) | 4 (100%) | 15 (88%) |
| Romanello et al. [37] | 4 (100%) | 3 (75%) | 5 (100%) | 4 (100%) | 16 (94%) |
| Ruiz-Rizzo et al. [38] | 4 (100%) | 3 (75%) | 5 (100%) | 4 (100%) | 16 (94%) |
| Saberi et al. [39] | 4 (100%) | 3 (75%) | 4 (80%) | 4 (100%) | 15 (88%) |
| Specogna et al. [40] | 3 (75%) | 3 (75%) | 5 (100%) | 4 (100%) | 15 (88%) |
| Štecková et al. [41] | 4 (100%) | 3 (75%) | 5 (100%) | 4 (100%) | 16 (94%) |
| Svolgaard et al. [42] | 4 (100%) | 3 (75%) | 5 (100%) | 3 (75%) | 14 (82%) |

| | | | | | |
|---|---|---|---|---|---|
| Svolgaard et al. [43] | 4 (100%) | 3 (75%) | 5 (100%) | 4 (100%) | 16 (94%) |
| Téllez et al. [44] | 3 (75%) | 3 (75%) | 5 (100%) | 4 (100%) | 15 (88%) |
| Tomasevic et al. [45] | 3 (75%) | 3 (75%) | 5 (100%) | 4 (100%) | 15 (88%) |
| Wilting et al. [46] | 3 (75%) | 3 (75%) | 5 (100%) | 4 (100%) | 15 (88%) |
| Wu et al. [47] | 4 (100%) | 3 (75%) | 5 (100%) | 4 (100%) | 16 (94%) |
| Wu et al. [48] | 4 (100%) | 3 (75%) | 5 (100%) | 4 (100%) | 16 (94%) |
| Yaldizli et al. [49] | 4 (100%) | 3 (75%) | 5 (100%) | 4 (100%) | 16 (94%) |
| Zellini et al. [50] | 3 (75%) | 1 (25%) | 4 (80%) | 4 (100%) | 12 (71%) |
| Zhou et al. [51] | 4 (100%) | 3 (75%) | 5 (100%) | 4 (100%) | 16 (94%) |
| Zhou et al. [52] | 4 (100%) | 3 (75%) | 5 (100%) | 4 (100%) | 16 (94%) |
