## Supplemental Table 4 for "Brain connectivity changes underlying depression and fatigue in relapsing-remitting multiple sclerosis: a systematic review"

**S4 Table. Quality assessment of longitudinal studies using the Institute of Health Economics (IHE) 'Quality Appraisal of Case Series Studies Checklist' [1].** The authors of this review assigned values to the checklist answers. Percentages are based on the number of 'positively' answered questions per category, with 15 questions in total. Irrelevant questions were excluded. A high percentage indicates higher quality.

| **Author** | **Study objective (1 max)** | **Study design (3 max)** | **Study population (3 max)** | **Outcome measure (2 max)** | **Statistical analysis (1 max)** | **Results and conclusions (4 max)** | **Competing interests and sources of support (1max)** | **Total (15 max)** |
|---|---|---|---|---|---|---|---|---|
| Calabrese et al. [54] | 1(100%) | 2(67%) | 1.5(50%) | 2(100%) | 1(100%) | 3(75%) | 1(100%) | 10.5(70%) |
| Cavallari et al. [55] | 1(100%) | 1(33%) | 2(67%) | 2(100%) | 1(100%) | 4(100%) | 1(100%) | 11(73%) |
| Hildebrandt and Eling [56] | 0.5(50%) | 1(33%) | 2(67%) | 2(100%) | 0(0%) | 2(50%) | 1(100%) | 7.5(50%) |
| Yaldizli et al. [57] | 1(100%) | 2(67%) | 2(67%) | 2(100%) | 1(100%) | 3(75%) | 1(100%) | 11(73%) |
| Yarraguntla et al. [58] | 1(100%) | 1(33%) | 3(100%) | 2(100%) | 1(100%) | 3(75%) | 1(100%) | 11(73%) |
| Yarraguntla et al. [59] | 1(100%) | 2(67%) | 2(67%) | 2(100%) | 1(100%) | 3.5(88%) | 0(0%) | 11.5(77%) |
| Gilio et al. [60] | 1(100%) | 1(33%) | 3(100%) | 1(50%) | 1(100%) | 3.5(88%) | 1(100%) | 10.5(70%) |
| Tijhuis et al. [61] | 1(100%) | 1(33%) | 2(67%) | 2(100%) | 1(100%) | 4(100%) | 1(100%) | 11(73%) |
