## Supplemental Table 5 for "Brain connectivity changes underlying depression and fatigue in relapsing-remitting multiple sclerosis: a systematic review"

**S5 Table. Full breakdown of AXIS scores [1].** ns = not stated, considered as a negative score.

| Study | Study design | | | | Selection bias | | | | Comparability/Method | | | | | Outcomes | | | | Total | |
|---|---|---|---|---|---|---|---|---|---|---|---|---|---|---|---|---|---|---|---|
| | Were the aims/objectives of the study clear? | Was the study design appropriate for the stated aim(s)? | Was the sample size justified (based on pre-study considerations of statistical power)? | Was the target/reference population clearly defined? (Is it clear who the research was about?) | Was the sample frame taken from an appropriate population base so that it closely represented the target/reference population under investigation? | Was the selection process likely to select subjects/participants that were representative of the target/reference population under investigation? | Was ethical aspect approval or consent of participants attained? | Were the risk factor and outcome variable measured appropriate to the aims of the study? | Were the risk factor and outcome variables measured correctly using instruments/ measurements been trialled, piloted or published previously? | Is it clear what was used to determined statistical significance and/or precisions estimates? (e.g. p-values, confidence intervals) | Were the methods (including statistical methods) sufficiently described to enable them to be repeated? | Are the basic data adequately described? | Were the results internally consistent? | Were the results presented for all the analyses described in the method? | Were the author's discussions and conclusions justified by the results? | Were the limitations of the study discussed? | Were there any funding sources or conflicts of interest that may affect the authors' interpretation of the results? | Score | Percentage |
| Alshehri et al. [2] | Y | Y | N | Y | Y | Y | Y | Y | Y | Y | Y | Y | Y | Y | Y | Y | N | 16 | 94 |
| Altermatt et al. [3] | Y | Y | N | Y | Y | Y | Y | Y | Y | Y | Y | Y | Y | Y | Y | Y | N | 16 | 94 |
| Andreasen et al. [4] | Y | Y | N | Y | Y | Y | Y | Y | Y | Y | Y | Y | Y | Y | Y | Y | N | 16 | 94 |
| Bauer et al. [5] | Y | Y | N | Y | N | Y | Y | Y | Y | Y | Y | Y | Y | Y | Y | Y | N | 15 | 88 |
| Beaudoin et al. [6] | Y | Y | N | Y | Y | Y | Y | Y | Y | Y | Y | Y | Y | Y | Y | Y | N | 16 | 94 |
| Benesova et al. [7] | Y | Y | N | Y | Y | Y | N | Y | Y | Y | Y | N | Y | Y | Y | N | ns | 12 | 71 |
| Bisecco et al. [8] | Y | Y | N | Y | Y | Y | Y | Y | Y | Y | Y | Y | Y | Y | Y | Y | N | 16 | 94 |
| Carotenuto et al. [9] | Y | Y | N | Y | Y | Y | Y | Y | Y | Y | Y | Y | Y | Y | Y | Y | N | 16 | 94 |
| Codella et al. [10] | Y | Y | N | Y | Y | Y | N | Y | Y | Y | Y | Y | Y | Y | Y | N | ns | 13 | 76 |
| Damasceno et al. [11] | Y | Y | N | Y | Y | Y | Y | Y | Y | Y | Y | Y | Y | Y | N | Y | N | 15 | 88 |
| Filippi et al. [12] | Y | Y | N | Y | Y | Y | Y | Y | Y | Y | Y | Y | Y | Y | Y | N | N | 15 | 88 |
| Finke et al. [13] | Y | Y | N | Y | Y | Y | Y | Y | Y | Y | Y | Y | Y | Y | Y | N | N | 15 | 88 |
| Gold et al. [14] | Y | y | N | Y | Y | Y | Y | Y | Y | Y | Y | Y | Y | Y | Y | Y | N | 16 | 94 |
| Golde et al. [15] | Y | Y | Y | Y | N | Y | Y | Y | Y | Y | Y | Y | Y | Y | Y | Y | N | 16 | 94 |
| Cruz Gomez et al. [16] | Y | Y | Y | Y | N | Y | Y | Y | Y | N | Y | Y | Y | Y | Y | Y | N | 15 | 88 |

| | | | | | | | | | | | | | | | | | | |
|---|---|---|---|---|---|---|---|---|---|---|---|---|---|---|---|---|---|---|
| Hassan et al. [17] | Y | Y | Y | Y | N | Y | Y | Y | Y | N | Y | N | N | Y | Y | Y | N | 13 | 76 |
| Hildebrandt et al. [18] | Y | Y | N | Y | Y | Y | Y | Y | Y | Y | Y | Y | Y | Y | Y | Y | ns | 15 | 88 |
| Huang et al. [19] | Y | Y | N | Y | Y | Y | Y | Y | Y | Y | Y | Y | Y | Y | Y | Y | N | 16 | 94 |
| Iancheva et al. [20] | Y | Y | N | Y | Y | Y | Y | Y | Y | Y | Y | N | Y | N | Y | Y | N | 14 | 82 |
| Jaeger et al. [21] | Y | Y | N | Y | Y | Y | Y | Y | Y | Y | Y | Y | Y | Y | Y | Y | N | 16 | 94 |
| Kever et al. [22] | Y | Y | N | Y | Y | Y | Y | Y | Y | Y | Y | N | Y | Y | Y | Y | N | 15 | 88 |
| Khedr et al. [23] | Y | Y | N | Y | Y | Y | Y | Y | Y | Y | Y | Y | Y | Y | Y | Y | N | 16 | 94 |
| Kopchak and Odintsova [22] | Y | Y | N | Y | Y | Y | Y | N | N | Y | N | N | Y | Y | N | N | N | 10 | 59 |
| Lazzarotto et al. [24] | Y | Y | N | Y | Y | Y | Y | Y | Y | Y | Y | Y | N | Y | Y | Y | N | 15 | 88 |
| Morgante et al. [25] | Y | Y | N | Y | Y | Y | Y | Y | Y | Y | Y | Y | Y | Y | Y | Y | N | 16 | 94 |
| Niepel et al. [26] | Y | Y | N | Y | Y | Y | Y | Y | Y | Y | Y | Y | Y | Y | Y | N | ns | 14 | 82 |
| Nigro et al. [27] | Y | Y | N | Y | Y | Y | Y | Y | Y | Y | Y | Y | Y | Y | Y | Y | N | 16 | 94 |
| Nygaard et al. [28] | Y | Y | N | Y | Y | Y | Y | Y | Y | Y | Y | Y | Y | Y | Y | Y | N | 16 | 94 |
| Pardini et al. [29] | Y | Y | N | Y | Y | Y | Y | Y | Y | Y | Y | Y | Y | Y | Y | N | ns | 14 | 82 |
| Pardini et al. [30] | Y | Y | N | Y | Y | Y | ns | Y | Y | N | Y | N | Y | Y | Y | Y | N | 13 | 76 |
| Pokryszko-Dragan et al. [31] | Y | Y | N | Y | Y | Y | Y | Y | Y | Y | Y | Y | Y | Y | Y | Y | N | 16 | 94 |
| Pravatà et al. [32] | Y | Y | N | Y | Y | Y | Y | Y | Y | Y | Y | Y | Y | Y | Y | Y | N | 16 | 94 |
| Riccelli et al. [33] | Y | Y | N | Y | Y | Y | Y | Y | Y | Y | Y | Y | Y | Y | Y | Y | N | 16 | 94 |
| Rocca et al. [34] | Y | Y | N | Y | Y | Y | Y | Y | Y | Y | Y | Y | Y | Y | Y | Y | N | 16 | 94 |
| Rocca et al. [35] | Y | Y | N | Y | Y | Y | Y | Y | Y | Y | Y | Y | Y | Y | Y | Y | N | 16 | 94 |
| Rojas et al. [36] | Y | Y | N | Y | Y | Y | Y | Y | Y | Y | Y | Y | Y | Y | Y | N | N | 15 | 88 |
| Romanello et al. [37] | Y | Y | N | Y | Y | Y | Y | Y | Y | Y | Y | Y | Y | Y | Y | Y | N | 16 | 94 |
| Ruiz-Rizzo et al. [38] | Y | Y | N | Y | Y | Y | Y | Y | Y | Y | Y | Y | Y | Y | Y | Y | N | 16 | 94 |
| Saberi et al. [39] | Y | Y | N | Y | Y | Y | Y | Y | Y | Y | Y | N | Y | Y | Y | Y | N | 15 | 88 |
| Specogna et al. [40] | Y | Y | N | Y | Y | Y | Y | Y | Y | Y | Y | Y | Y | Y | Y | N | N | 15 | 88 |
| Štecková et al. [41] | Y | Y | N | Y | Y | Y | Y | Y | Y | Y | Y | Y | Y | Y | Y | Y | N | 16 | 94 |
| Svolgaard et al. [42] | Y | Y | N | Y | N | Y | Y | Y | Y | Y | Y | Y | Y | Y | Y | Y | ns | 14 | 82 |
| Svolgaard et al. [43] | N | Y | N | Y | Y | Y | Y | Y | Y | Y | Y | Y | Y | Y | Y | Y | N | 16 | 94 |
| Téllez et al. [44] | Y | Y | N | Y | Y | Y | Y | Y | Y | Y | Y | Y | Y | Y | Y | N | N | 15 | 88 |
| Tomasevic et al. [45] | Y | Y | N | Y | Y | Y | Y | Y | Y | Y | Y | Y | Y | Y | Y | N | N | 15 | 88 |
| Wilting et al. [46] | Y | Y | N | Y | Y | Y | Y | Y | Y | Y | Y | Y | Y | Y | Y | N | N | 15 | 88 |
| Wu et al. [47] | Y | Y | N | Y | Y | Y | Y | Y | Y | Y | Y | Y | Y | Y | Y | Y | N | 16 | 94 |
| Wu et al. [48] | Y | Y | Y | Y | N | Y | Y | Y | Y | Y | Y | Y | Y | Y | Y | Y | N | 16 | 94 |
| Yaldizli et al. [49] | Y | Y | N | y | Y | Y | Y | Y | Y | Y | Y | Y | Y | Y | Y | Y | N | 16 | 94 |
| Zellini et al. [50] | Y | Y | N | N | Y | N | Y | N | Y | Y | Y | Y | Y | Y | Y | N | N | 12 | 71 |

| | | | | | | | | | | | | | | | | | | |
|---|---|---|---|---|---|---|---|---|---|---|---|---|---|---|---|---|---|---|
| Zhou et al. [51] | Y | Y | Y | Y | N | Y | Y | Y | Y | Y | Y | Y | Y | Y | Y | Y | N | 16 | 94 |
| Zhou et al. [52] | Y | Y | N | Y | Y | Y | Y | Y | Y | Y | Y | Y | Y | Y | Y | Y | N | 16 | 94 |
