## Supplemental Table 6 for "Brain connectivity changes underlying depression and fatigue in relapsing-remitting multiple sclerosis: a systematic review"

**S6 Table. Full breakdown of Institute of Health Economics (IHE) checklist scores [1].** Y=Yes; N=No; ?=Unclear; P=Partial; Y*=retrospective study, same points as a 'yes' answer.

| | Study objective | Study design | | | Study population | | | Outcome measure | | Statistical analysis | Results and conclusions | | | | Competing interests and sources of support | Total |
|---|---|---|---|---|---|---|---|---|---|---|---|---|---|---|---|---|
| Paper | Was the hypothesis/aim/objective of the study clearly stated? | Was the study conducted prospectively? | Were the cases collected in more than one center? | Were patients recruited consecutively? | Were the characteristics of the patients included in the study described? | Were the eligibility criteria (i.e. inclusion and exclusion criteria) for entry into the study clearly stated? Were the eligibility criteria (i.e. inclusion and exclusion criteria) for entry into the study clearly stated? | Did patients enter the study at a similar point in the disease? | Were relevant outcome measures established a priori? | Were the relevant outcomes measured using appropriate objective/ subjective methods? | Were the statistical tests used to assess the relevant outcomes appropriate? | Was follow-up long enough for important events and outcomes to occur? | Were losses to follow-up reported? | Did the study provided estimates of random variability in the data analysis of relevant outcomes? | Were the conclusions of the study supported by the results? | Were both competing interests and sources of support for the study reported? | |
| Calabrese et al. [2] | Y | Y | N | Y | Y | P | N | Y | y | y | Y | N | Y | y | y | 10.5 (70%) |
| Cavallari et al. [3] | Y | Y | ? | ? | Y | Y | N | Y | Y | Y | Y | Y* | Y | Y | Y | 11 (73%) |
| Hildebrandt and Eling [4] | P | Y | ? | ? | Y | Y | N | Y | Y | ? | Y | N | Y | ? | Y | 7.5 (50%) |
| Yaldizli et al. [5] | Y | Y | N | Y | Y | Y | N | Y | Y | Y | Y | N | Y | Y | Y | 11 (73%) |
| Yarraguntla et al. [6] | Y | Y | N | ? | Y | Y | Y | Y | Y | Y | Y | N | Y | Y | Y | 11 (73%) |
| Yarraguntla et al. [7] | Y | N | N | ? | Y | Y | N | Y | y | Y | Y | Y* | P | Y | N | 11.5 (77%) |
| Gilio et al. [8] | Y | N | N | Y | Y | Y | Y | ? | Y | Y | Y | Y* | P | Y | Y | 10.5 (70%) |
| Tijhuis et al. [9] | Y | Y | ? | ? | Y | Y | N | Y | Y | Y | Y | Y | Y | Y | Y | 11 (73%) |
