## Supplemental Table 7 for "Brain connectivity changes underlying depression and fatigue in relapsing-remitting multiple sclerosis: a systematic review"

**S7 Table. Overview of study details for publications included (N=60) in the current systematic review.**

| Authors | MRI sequence | Field Strength | Design | Participants (n) | Sex: no. female (%) | Country | Source of participants |
|---|---|---|---|---|---|---|---|
| Alshehri et al. [1] | T1W, FLAIR, DTI | 3T | Cross-sectional | RRMS: 37; HC: 19;<br>EDSS ≤ 4.0 (1.9±0.15) | RRMS 37(78%)<br>HC 19(80%) | Australia | Patients |
| Altermatt et al. [2] | T1W, T2W, FLAIR | 1T/1.5T/3T | Retrospective | RRMS (1907)<br>Mean age: 39<br>EDSS: 2 | RRMS 1362 (71%) | Multicentre, mostly the United States) | All available data from patients participating in the multicentre FREEDOMS (N = 1272) and FREEDOMS II (n = 1083) phase 3 clinical trials |
| Andreasen et al. [3] | T1W, T2W, FLAIR, DTI, MRS proton spectroscopy | 3T | Cross-sectional | RRMS (34): F (17), nF (17)<br>HC (7)<br>Mean age: 43 (F), 39 (nF), 39 (HC)<br>EDSS: 1-3.5 | F 12(71%)<br>nF 8(47%)<br>HC 6(86%) | Denmark | Patients |
| Bauer et al. [4] | T1W, T2W, FLAIR, DWI (dMRI) | 3T | Cross-sectional | RRMS (46); HC (25);<br>Mean age(range)[DS]: RRMS: 36.1(22–53)[8.6]; HC: 35.8 (19–55)[10.6]; EDSS Mean 1.5(0–3.5)[1.3] | MS: 33 (72%)<br>HC: 16(64%) | Denmark | Patients |
| Beaudoin et al. [5] | T1W, T2W, HARDI | 3T | Cross-sectional | RRMS:24; HC: 11;<br>EDSS(median[range]): 1.5[0-3] | RRMS 20(83%)<br>HC: 8(73%) | Canada | Patients |
| Benesova et al. [6] | T1W, T2W | 1.5T | Cross-sectional | RRMS (20): D (10), nD (10);<br>EDSS: 1-4<br>Mean age: 37.3 | RRMS 14(70%) | Czech Republic | Patients |
| Bisecco et al. [7] | T1W, T2W, DTI | 3T | Cross sectional | RRMS (60): F (30), nF (30), HC (29);<br>Mean age: 40.2 (nF), 40.7 (F)<br>EDSS: 2(F), 1.5 (nF) | RRMS 41(68%)<br>nF 21(70%)<br>F 20(67%)<br>HC 16(55%) | Italy | Patients. HC were recruited from a large HC database. |
| Calabrese et al. [8] | FLAIR, T2W | 1.5T | Longitudinal | RRMS (152)<br>HC (42)<br>Mean age: 34 (RRMS), 35.5 (HC)<br>EDSS: 2.7 | RRMS 95(63%)<br>HC 26(62%) | Italy | Patients |
| Carotenuto et al. [9] | T1, T2, FLAIR, rs-fMRI | 3T | Cross-sectional | RRMS (29), HC (24)<br>Mean age: 42 (RRMS), 38 (HC)<br>Mean EDSS: 3.2 | RRMS 17(59%)<br>HC 15(63%) | UK | Patients |
| Cavallari et al. [10] | T2W | 1.5T | Retrospective | RRMS (66)<br>Mean age: 48<br>EDSS: 1.5 | RRMS 52(79%)<br>Converters* 26(79%)<br>Non-converters 26(79%) | USA | Retrospectively selected from a larger cohort of over 800 prospectively followed MS patients within the CLIMB study |
| Codella et al. [11] | DE TSE, 2D GE, pulsed gradient spin-echo echo-planar | 1.5T | Cross-sectional | RRMS (28): F (14), nF (14)<br>HC (30)<br>Mean age: 37.6 (nF), 39.1 (F)<br>Median EDSS: 1 | RRMS 19(68%)<br>HC 18(60%) | Italy | Patients |
| Damasceno et al. [12] | FLAIR, T1W, T2W | 3T | Cross-sectional | RRMS (49)<br>HC (30)<br>Mean age: 30.94 (RRMS), 29.52 (HC) | RRMS 38 (77.6%)<br>HC 23 (76.7%) | Brazil | Patients |

| | | | | | | | |
|---|---|---|---|---|---|---|---|
| | | | | Median EDSS: 2 | | | |
| Filippi et al. [13] | T1W, fMRI, DE TSE | 1.5T | Cross-sectional | RRMS (29): F (15), nF (14), HC (15);<br>Mean age: 39.3 (F), 37.6 (nF)<br>Median EDSS: 1 | MS 20(69%)<br>HC 9(60%) | Italy | Patients |
| Finke et al. [14] | T2W, DTI, T1, FLAIR, rs-fMRI | 3T | Cross-sectional | RRMS (44)<br>HC (20)<br>Mean age: 45.9 (RRMS), 43.1 (HC) | MS 26(59%)<br>HC 11(55%) | Germany | Patients |
| Gilio et al. [15] | T1W, FLAIR, PD, FLAIR, Gd+ T1W | 3T | Longitudinal | RRMS: 106 (35 had MRI);<br>EDSS(median[IQR]): 1.5 [1–2.125] | RRMS 106 (66%) | Italy | patients |
| Gold et al. [16] | T1W, T2W, FLAIR | 3T | Cross-sectional | RRMS (29): D (21), nD (8)<br>HC (20)<br>Mean age: 37.5<br>Mean EDSS: 2.5 | RRMS 25(86%)<br>HC 18(90%) | USA | Patients |
| Golde et al. [17] | BOLD, MP-RAGE, DTI | 3T | Cross-sectional | RRMS (30), HC (34);<br>Age: RRMS 40.20±9.87; HC 39.57±8.36;<br>EDSS: 1.75 (0–4) | RRMS: 18(60%);<br>HC: 19(63%) | Germany | Patients |
| Gómez et at. [18] | T1W, T2*W | 1.5T | Cross-sectional | RRMS (60), nF (28), F(32);<br>HC (18);<br>Mean age(SD)[range]:<br>HC 31.06(5.67)[22-44];<br>nF: 34.96(5.87)[20-44];<br>F: 37.72(5.90)[22-47];<br>EDSS mean(SD)[range]<br>nF: 1.96(1.20)[0-5];<br>F 3.20(1.68)[1-6] | HC: 8(44%);<br>nF: 18(64%);<br>F: 21(66%) | Spain | Patients |
| Hassan et al. [19] | T1W, T2W, FLAIR, DWI | 1.5T | Cross-sectional | D-RRMS (20); nD-MS (10);<br>HC (10);<br>Mean age (range): 27 (21-36);<br>EDSS ≤ 5 | MS: 24(80%) | Egypt | Patients |
| Hildebrandt et al. [20] | T1W | 1.5T | Cross-sectional | RRMS (45)<br>Mean age: 38.9<br>Median EDSS: 2.6 | RRMS 29(64%) | Germany | Patients |
| Hildebrandth and Eling [21] | T1W | 1.5T | Longitudinal | 40 RRMS: no increase in F (23),<br>increase in F (17);<br>Mean age: 38.5 (CIF), 37.9 (NCIF)<br>EDSS: 2.3 (CIF), 3.2 (NCIF) | No increase in F 17(74%)<br>Increase in F (9(53%) | Germany | Patients |
| Huang et al. [22] | T1W, T2W, FLAIR, rs-fMRI | 3T | Cross-sectional | RRMS (33)<br>HC (33)<br>Mean age: 41.8 (RRMS), 42.2 (HC)<br>EDSS: 1.97 | RRMS 21(64%)<br>HC 21(64%) | China | Patients |
| Iancheva et al. [23] | T1, fMRI | 3T | Cross sectional | RRMS (29): F (15), nF (14)<br>Mean age: 40.7 (F), 36.9 (nF)<br>Median EDSS:1 | No data | Bulgaria | Patients |

| | | | | | | | |
|---|---|---|---|---|---|---|---|
| Jaeger et al. [24] | T1W, T2W, rs-fMRI | 3T | Cross sectional | RRMS (70): F (39), nF (38)<br>Median age: 40 (F), 34.5 (nF), 36 (HC)<br>Median EDSS: 2.5 (F), 2 (nF) | F 32(82%)<br>nF 24(63%)<br>HC 26(63%) | Germany | Recruited from ongoing prospective studies |
| Kever et al. [25] | T1W, FLAIR, T2W | 3T | Longitudinal | RRMS: 51;<br>EDSS(median(IQR)): 1.5 (4.5) | RRMS (80.4%) | USA | MEM CONNECT cohort |
| Khedr et al. [26] | PD, FLAIR, T1W, T2W, Gd+T1W | 1.5T | Cross-sectional | 43 RRMS patients with 40 ;<br>EDSS (mean±SD) F: 4.13±1.59;<br>nF: 2.42±1.17 | RRMS:<br>F 24(77%)<br>nF 6(50%) | Egypt | Patients |
| Kopchak & Odintsova [27] | ? | 1.5T | Cross-sectional | RRMS: 106; A group: under 40 years (n=48); B group: —≥40 (n=58); EDSS score: 6.5—8 points (EXCLUSION) | RRMS 81(76%) | Ukraine | Patients |
| Lazzarotto et al. [28] | T1W, FLAIR, DIR | 3T | Cross-sectional | RRMS (61)<br>HC (56)<br>Mean age:<br>37.9 (RRMS),<br>35.2 (HC)<br>EDSS: 1.77 | RRMS 43(70%)<br>HC 34(61%) | Italy | Retrospective: Data were retrieved from the MRI and clinical databases of the MS center of Padua, screening all patients that came to our attention from April 2014 to May 2018 |
| Morgante et al. [29] | T1 GD+, T2W, transcranial magnetic stimulation | 1.5T | Cross sectional | RRMS (33): F (16), nF (17)<br>HC (12)<br>Mean age: 41 (F), 38 (nF),<br>EDSS: 1.8 (F), 1.6 (nF) | nF 13(77%)<br>F 9(56%) | Italy | Patients |
| Niepel et al. [30] | 3D FLASH (for T1maps), T2 | 1.5T | Cross-sectional | RRMS (52[48 for FSS]; 34 for MRI, clinical): F (39[20 for MRI]), nF (23[11 for MRI]). HC (19)<br>Mean age: 39 (F), 33 (nF)<br>Median EDSS: 3 (F), 2 (nF) | RRMS 40(77%)<br>[37(77%) for FSS; 27(79%) for MRI, clinical]<br>HC 14(74%) | UK | Patients |
| Nigro et al. [31] | T1W, FLAIR, DTI | 3T | Cross-sectional | RRMS (42): D (20), nD (22). HC (16)<br>Mean age: 37.1 (D), 31.5 (nD), 35.3 (HC)<br>EDSS: 2.3(nD), 2.8(D) | D (85%)<br>nD (60%)<br>HC (43%) | Italy | Patients |
| Nygaard et al. [32] | T1W, FLAIR | 1.5T | Cross-sectional | RRMS (61)<br>HC (61);<br>EDSS: 0-4;<br>Mean age: RRMS 34.2, HC 33.5 | RRMS 47(77%)<br>HC 47(77%) | Norway | Patients |
| Pardini et al. [33] | T1W, T2W, DTI | 1.5T | Cross-sectional | RRMS (40): F (15), nF (25), HC (15);<br>Mean age: 41 (F), 36 (nF)<br>Mean EDSS: 1.6 | RRMS 28(70%)<br>HC no data | Italy | Patients |
| Pardini et al. [34] | T1W, T2W, DTI | 1.5T | Cross-sectional | RRMS (77): F (25), nF (52)<br>Mean age: 40.8<br>EDSS: 2 | No data | Italy | Patients<br>(Reporting fatigue) |
| Pokryszko-Dragan et al. [35] | T1W, T2W, FLAIR, DWI, DTI, 3D-FSPGR GD+ | 1.5T | Cross-sectional | RRMS (50)<br>HC (27)<br>Mean age: 36.4 (RRMS), 36.3 (HC)<br>EDSS: 2.7 | RRMS 37(74%)<br>HC 19(70%) | Poland | Patients |

| | | | | | | | |
|---|---|---|---|---|---|---|---|
| Pravatà et al. [36] | T1W, T2W, rs-fMRI | 3T | Cross-sectional | RRMS (22): F (11), nF (11)<br>HC (12)<br>Mean age: 46.6 (F), 40 (nF), 41.4 (HC)<br>EDSS: 1.5 (nF), 2.5 (F) | HC 6(50%)<br>nF 4(57%)<br>F 4(57%) | Switzerland | Patients |
| Riccelli et al. [37] | T1W, FLAIR, fMRI | 3T | Cross sectional | RRMS (77),<br>HC (20)<br>Mean age: 34 (RRMS), 36.4 (HC)<br>Median EDSS: 2 | RRMS 46(60%)<br>HC 10(50%) | UK | Patients |
| Rocca et al. [38] | T2W, T1W, fMRI | 1.5T | Cross sectional | RRMS (24): F (11), nF (13)<br>HC (46)<br>Mean age: 33.8 (F), 31.2 (nF)<br>Median EDSS: 1 (both groups) | RRMS 22(92%) | Italy | Patients |
| Rocca et al. [39] | T2W, T1W, fMRI | 3T | Cross-sectional | RRMS (79): F (50), nF (29)<br>HC (26)<br>Mean age: 42.6 (F), 40 (nF), 39.2 (HC)<br>Median EDSS: 2 (F), 1.5 (nF) | HC 17(65%)<br>nF-MS 19(66%)<br>F-MS 33(66%) | Italy | Patients |
| Rojas et al. [40] | T1W, T2W, FLAIR, DTI | 1.5T | Cross-sectional | RRMS (45): D (23), nD (22);<br>Mean age: 33.5 (D), 32.5 (nD), 32.3 (HC)<br>EDSS: 1.6 (nD), 2 (D) | HC 14(70%)<br>nD 14(63.6%)<br>D 19(82.6%) | Argentina | Patients |
| Romanello et al. [41] | T2W, rs-fMRI | 3T | Cross-sectional | RRMS: 101; HC: 101;<br>EDSS (median(IQR)) RRMS 1.5 (1.5);<br>(EDSS ≤ 1, n = 36) 1 (1); (EDSS ≥ 2, n = 39) 2.5 (1) | RRMS 67(66%)<br>(EDSS ≤ 1) 20(56%);<br>(EDSS ≥ 2) 27(69%); HC 67(66%) | Germany | Patients |
| Ruiz-Rizzo et al. [42] | T1W, T2*W, FLAIR, BOLD fMRI | 3T | Cross-sectional | RRMS: 104;<br>EDSS<3: 61 (58.7%); 3 – 7: 43 (41.3%) | RRMS 66(63.5%) | Germany | Patients |
| Saberi et al. [43] | T1W | 1.5T | Cross sectional | RRMS (43)<br>EDSS ≤ 6<br>Age 21-59 | RRMS 32 (74%) | Iran | Recruited from the Cross-Modal Research Initiative for MS and Optic Neuritis (CRIMSON) |
| Specogna et al. [44] | T1W, T2W, T1 GD+, FLAIR, fMRI: finger against thumb tapping | 1.5T | Cross sectional | RRMS (24): F (12), nF (12)<br>HC (15)<br>Mean age: 39.8 (F), 38.7 (nF)<br>EDSS: 1.5 | RRMS 20(83%) | Italy | Patients |
| Štecková et al. [45] | T1W | 1.5T | Cross-sectional | RRMS (43)<br>HC (19)<br>Mean age: 35.2 (MS5), 43.5 (MS10)<br>EDSS: not stated | RRMS 29(67%)<br>CIS 12(63%)<br>MS5 11(73%)<br>MS10 6(67%) | Czech Republic | Patients |
| Svolgaard et al. [46] | T1W, T2W, FLAIR, fMRI | 3T | Cross-sectional | RRMS (24): F (12), nF (12)<br>HC (15)<br>Mean age: 40.9 (F), 38.7 (nF)<br>EDSS: 1.5 | MS 30(68%)<br>HC 16(64%) | Germany | No information |
| Svolgaard et al. [47] | T1W, T2W, FLAIR, fMRI | 3T | Cross-sectional | RRMS: 44; HC: 25;<br>EDSS score ≤ 3.5 | RRMS 30(68%)<br>HC 16(64%) | Denmark | Patients |

| | | | | | | | |
|---|---|---|---|---|---|---|---|
| Tellez et al. [48] | T2, proton magnetic resonance | 1.5T | Cross sectional | RRMS (30): F (17), nF (13)<br>HC (21)<br>Mean age: 38.5 (F), 37.8 (nF)<br>Median EDSS: 2.5 (F), 1.5 (nF) | F 13(76.4%)<br>nF 10(78.1%) | Spain | Patients |
| Tijhuis et al. [49] | T1W, FLAIR, fMRI | 3T | Longitudinal | RRMS: 35; HC: 19;<br>EDSS Baseline 3(1–6)<br>EDSS Follow up 3(1.50–7) | RRMS 20(57%)<br>HC 11(58%) | The Netherlands | Patients |
| Tomasevic et al. [50] | T1 GD+, T2, T1 FLAIR | 1.5T | Cross-sectional | RRMS (20): F (11), nF (9)<br>Mean age: 38.5 (F); 35.9 (nF)<br>EDSS: 0-2 | RRMS 13(65%)<br>nF 6(67%)<br>F 8(73%) | Italy | Patients |
| Wilting et al. [51] | T1W, FLAIR, DTI | 3T | Cross-sectional | RRMS (79): F (38), nF (41)<br>Median age: 34.5 (F), 30 (nF)<br>Median EDSS: 2 | nF 26(63%)<br>F 30(79%)<br>HC 18(45%) | Germany | Patients |
| Wu et al. [52] | T1W, T2W, rs-fMRI | 3T | Cross-sectional | RRMS (22)<br>HC (22)<br>Mean age: 44.6 (RRMS), 40.1 (HC)<br>EDSS: 1.8 | RRMS 13(59%)<br>HC 13(59%) | China | Patients |
| Wu et al. [53] | T2*W, T2W, T1W, rs-fMRI | 3T | Cross-sectional | RRMS (41), HC (23);<br>Mean age(range): acute RRMS: 43.1 (15–61), remitting RRMS: 40.7 (21–66), HC 40.7 (26–58);<br>Mean EDSS (range):<br>acute RRMS 2.8 (1.5–4);<br>remitting RRMS 2.0 (0–3.5) | RRMS:<br>acute: 10(59%);<br>remitting 15(63%); HC 11(49%); | China | Patients |
| Yaldizli et al. [54] | T1 GD+, T1W, T2W, FLAIR | 1.5T | Retrospective | RRMS (70): F (28), nF (42). HC (27)<br>Mean age: 40.4 (RRMS), 43.7 (HC)<br>EDSS: 2.8 | Total RRMS 61(67.1%)<br>FSS<4 37(88.1%)<br>FSS≥24(85.7%) | Switzerland | Patients |
| Yaldizli et al. [55] | T1W, PD/T2W | 1.5T | Cross-sectional | RRMS (103)<br>Mean age: 49<br>Median EDSS: 3 | RRMS 96(65.8%)<br>RRMS 73(70.9%)<br>SPMS 18(51.4%)<br>PPMS 5(62.5%)<br>HC 17(63%) | Switzerland | Recruited from an ongoing prospective, non-interventional cohort study on the phenotype-genotype characterization of MS |
| Yarraguntla et al. [56] | T1W, DTI | 3T | Retrospective longitudinal | RRMS (43): HF (15), MF (14), LF (14)<br>Mean age: 43 (HF), 39 (MF), 39 (LF)<br>EDSS: 3.6 (HF), 2.7 (MF), 2.35 LF) | No data | USA? | Retrospective longitudinal study from the Wayne State University School of Medicine MS Center |
| Yarraguntla et al. [57] | T2, FLAIR, MR spectroscopy | 3T | Longitudinal, 1 year | RRMS (48): HF (16), MF (18), LF (14)<br>Mean age: 43 (HF), 39 (MF), 39 (LF)<br>Median EDSS: 3 (HF), 2.71 (MF), 2.42 (LF) | RRMS 33(69%)<br>HF 12(75%)<br>MF 14(78%)<br>LF 7(50%) | USA | Patients |
| Zellini et al. [58] | T1W, T2W | 1.5T | Cross sectional | RRMS (44[40 for F tests, 36 intermediate count, 32 final count),<br>HC (13)<br>Median age: 39<br>Median EDSS: 2.5 | RRMS 35(80%)<br>[29(81%)]<br>HC 9(69%) | UK | Patients |

| | | | | | | | |
|---|---|---|---|---|---|---|---|
| Zhou et al. [59] | T2W, T1W, DTI, rs-fMRI | 3T | Cross-sectional | RRMS (24)<br>HC (24) | RRMS: 16(67%)<br>HC: 16(67%) | China | Patients |
| Zhou et al. [60] | T1W, T2W, DTI, rs-fMRI, FLAIR | 3T | Cross sectional | RRMS (20)<br>HC (20)<br>Mean age: 39.4 (RRMS), 38.1 (HC)<br>EDSS: 1.67 | RRMS 15(75%)<br>HC 15(75%) | China | Patients |

BOLD: Blood-Oxygen Level Dependent, D/nD: [not]depression/depressed, DTI: Diffusion Tensor Imaging, EDSS: Expanded Disability Status Scale, F/nF: [not]fatigue/fatigued, FA: Fractional Anisotropy, FLAIR: Fluid-attenuated Inversion Recovery, fMRI: functional Magnetic Resonance Imaging, RRMS: Relapsing-remitting multiple sclerosis, rs-fMRI: Resting-state Functional Magnetic Resonance Imaging, SD: Standard Deviation, HC: healthy controls, GD+: Gadolinium Enhancing, SPMS: Secondary progressive MS, PPMS: Primary progressive MS, LF/MF/HF: low/medium/high fatigue, FSPGR: fast spoiled gradient echo, DLPFC: dorsolateral prefrontal cortex. *Converters: converted to confirmed (⩾2 years) EDSS score ⩾3 within a follow-up period ⩾3 years.
