## Supplemental Table 8 for "Brain connectivity changes underlying depression and fatigue in relapsing-remitting multiple sclerosis: a systematic review"

**S8 Table. Negative findings explicitly reported in the included studies of this systematic review.** As studies did not fully report on all observed null findings, this table serves as an approximate summary of negative findings.

| MRI | Area | Depression | Overlap | Fatigue |
|---|---|---|---|---|
| Conventional MRI | Amygdala | Riccelli et al. [1] | Riccelli et al. [1]; Kever et al. [2] | Kever et al. [2] |
| | Hippocampus | Riccelli et al. [1] | - | - |
| | Basal ganglia volumes | - | - | Finke et al. [3]; Codella et al. [4] |
| | CA1, Subiculum, Entorhinal Cortex | Gold et al. [5] | - | - |
| | Caudate nucleus, superior ventral striatum | - | - | Jaeger et al. [6] |
| | Cerebral cortex of the frontal lobe | - | - | Codella et al. [4] |
| | Cerebral volume | - | - | Pardini et al. [7] |
| | corpus callosum index | Benesova et al. [8] | Yaldizli et al. [9]; Benesova et al. [8] | Yaldizli et al. [9] |
| | Cortical area | Nygaard et al. [10] | - | - |
| | Global atrophy | Gold et al. [5] | - | - |
| | Global brain parenchymal fraction | - | - | Cavallari et al. [11]; Andreasen et al. [12] |
| | Global lesion volume | Gold et al. [5]; Riccelli et al. [1]; Beaudoin et al. [13]; Romanello et al. [14] | Zellini et al. [15]; Niepel et al. [16]; Yaldizli et al. [9]; Calabrese et al. [17]; Gold et al. [5]; Filippi et al. [18]; Rocca et al. [19]; Rocca et al. [20]; Specogna et al. [21]; Svolgaard et al. [22]; Pravatà et al. [23]; Pardini et al. [7]; Wilting et al. [24]; Andreasen et al. [12]; Bisecco et al. [25]; Riccelli et al. [1]; Beaudoin et al. [13]; Alshehri et al. [26]; Romanello et al. [14] | ellini et al. [15]; Niepel et al. [16]; Yaldizli et al. [9]; Calabrese et al. [17]; Yaldizli et al. [9]; Filippi et al. [18]; Rocca et al. [19]; Rocca et al. [20]; Specogna et al. [21]; Svolgaard et al. [22]; Pravatà et al. [23]; Pardini et al. [7]; Wilting et al. [24]; Andreasen et al. [12]; Bisecco et al. [25]; Ruiz-Rizzo et al .[27]; Beaudoin et al. [13]; Alshehri et al. [26]; Romanello et al. [14] |
| | GM density, WM integrity | - | - | Finke et al. [3] |

| | | | | |
|---|---|---|---|---|
| | Hypothalamic volume | - | - | Zellini et al. [15] |
| | Lesion occurrence | - | - | Bisecco et al. [25] |
| | NAWM | - | - | Andreasen et al. [12] |
| | Normalized brain volume | Riccelli et al. [1]; | Bisecco et al. [25]; Riccelli et al. [1]; Rocca et al. [19]; Filippi et al. [18]; Rocca et al. [20] | Bisecco et al. [25]; Rocca et al. [19]; Filippi et al. [18]; Rocca et al. [20] |
| | Olfactory bulb volume | Yaldizli et al. [28] | - | - |
| | Putamen and caudate | - | - | Niepel et al. [16] |
| | Striatum | - | - | Jaeger et al. [6] |
| | Subgenual cingulate cortex | Riccelli et al. [1] | - | - |
| | T1 lesion burden | Rojas et al. [29] | - | - |
| | Temporal lobe | Benesova et al. [8] | - | - |
| | Thalamus volume | - | - | Saberi et al. [30] |
| | Total GM volume | Nigro et al. [31] | Nigro et al. [31]; Bisecco et al. [25]; Riccelli et al. [1]; Rocca et al. [19] | Bisecco et al. [25]; Rocca et al. [19]; Tijhuis et al. [32] |
| | Total intracranial volume | - | - | Saberi et al. [30] |
| | Volume fraction | - | Hildebrandt et al. [33] | - |
| | WM fraction | - | - | Wilting et al. [24] |
| | WM volume | Roj; as et al. [29]; Riccelli et al. [1] | Rojas et al. [29]; Bisecco et al. [25]; Riccelli et al. [1]; Rocca et al. [19] | Bisecco et al. [25]; Tijhuis et al. [32]; Rocca et al. [19] |
| **Structural connectivity** | FA, MD, RD, and AD | - | - | Bisecco et al. [25] |
| | DTI metrics in total brain WM | - | - | Alshehri et al. [26] |
| | DTI metrics in WML | - | - | Alshehri et al. [26] |
| | Frontal cortex | - | - | Wilting et al. [24] |
| | Global FA | Rojas et al. [29] | - | - |
| | Global FA, MD, PD, RD | - | - | Finke et al. [3] |
| | Global MD | - | - | Yarraguntla et al. [34] |
| | Thalamus | - | - | Bisecco et al. [25] |

| | | | | |
|---|---|---|---|---|
| **Functional connectivity** | Caudate and putamen and the dlPFC | - | - | Jaeger et al. [6] |
| | Cortical clusters in the prefrontal, premotor, sensorimotor, parietal, occipital cortex, cerebellum, basal ganglia bilaterally, putamen as ROI | - | - | Svolgaard et al. [22] |
| | Hippocampus and the rest of the brain | - | - | Golde et al. [35] |
| AD: axial diffusivity; dlPFC: dorsolateral prefrontal cortex; DTI: diffusion tensor imaging; FA: fractional anisotropy; GM: gray matter; MD: mean diffusivity; MRI: magnetic resonance imaging; NAWM: normal appearing WM; RD: radial diffusivity; ROI: region of interest; WM: white matter; WML: white matter lesions. | | | | |
